# Revealing Gender Differences in Concussion Reporting: A Detailed Analysis of SCAT Assessment Self-Report Symptom Ratings

**DOI:** 10.1101/2025.02.17.25322175

**Authors:** Rachel Edelstein, Karen M. Schmidt, John Darrell Van Horn

## Abstract

Current concussion assessments used by the NCAA are typically applied across both male and female athletes to assess the effects of sports-related head impacts. However, increasing evidence suggests that female athletes exhibit different physiological and psychosocial responses to concussions compared to their male counterparts, raising concerns about the appropriateness of gender-blind concussion assessment. This study analyzes data from 1,021 NCAA athletes (379 females, 642 males) who completed the SCAT3 Symptom Severity Checklist post-concussion. A systematic use of multivariate statistical approaches including Exploratory Graph Analysis (EGA), Principal Component Analysis (PCA), Exploratory Factor Analysis (EFA), Linear Discriminant Analysis (LDA), and Rasch Partial Credit Modeling (PCM) was employed on this 22-item instrument to explore the latent factor structure and to detect assessment items sensitive to potential gender differences. Differential Item Functioning (DIF) analysis was used to investigate gender disparities in symptom reporting. Based on EGA and PCA, the SCAT3 exhibited a four-factor substructure, with an EFA accounting for 62.44% of the variance. LDA between males and females displayed a statistically significant difference in the multivariate distributions of male and female scores (χ^2^(22) = 130.56, p < .001), indicating that emotional and physical symptom items loaded negatively, whereas cognitive and sensory-based items loaded positively. A closer examination of each assessment item via Rasch analysis revealed three items having zero difference between males and females. In contrast, nine symptoms indicated males were more likely to report higher severity. However, males generally reported overall lower total symptom severity scores (M = 30.06, SD = 20.88) and males (M = 24.71, SD = 21.18), t(765.06) = 3.85, p < .001. These differences in how athletes of differing genders present symptoms post-concussion may indicate that 1) males may be more conservative in their reporting and only endorse symptoms when they are felt to be more intense, contributing to high scores on a smaller number of symptoms, whereas 2) females may emphasize emotional and physical symptoms more readily. The results of this examination suggest that attention to gender differences in concussion symptom reporting should be considered in making clinical recommendations on concussion recovery and return-to-play decision-making.

## Introduction

Sports-related concussions (SRC) represent a significant health concern for student-athletes regardless of age, sport, athletic division/conference/etc., or gender. The *National Collegiate Athletic Association (NCAA)* employs a broad, multifaceted approach to determining SRC, but one which relies heavily upon symptom self-reporting as a primary method of assessment (Broglio et al., 2014; Kroshus et al., 2021; Meehan et al., 2013; Dick, 2009; McCrory et al., 2012; Harmon et al., 2013). However, increasing evidence suggests that male and female athletes exhibit distinct physiological and psychosocial responses to concussions, raising concerns about the adequacy of the current gender-neutral diagnostic framework (Nowitzki & Grant, 2011; Gessel et al., 2007; Granito, 2002; Kieffer et al., 2021; Mihalik et al., 2009; O’Connor et al., 2017; Schatz et al., 2011; Wallace et al., 2017; Zuckerman et al., 2014). Despite this growing body of research, NCAA concussion assessments, such as the widely used *Sport Concussion Assessment Tool (SCAT)*, do not specifically account for gender differences in symptom reporting or recovery trajectories (Echemendia et al., 2017).

Concussion rates among NCAA student-athletes underscore the importance of refining diagnostic tools. Over 460,000 student-athletes participate in NCAA competitions each year, nearly half (43.7%) of whom are female (Garcia et al., 2020; Musko & Demetriades, 2023). For instance, between the 2009–2010 and 2013– 2014 academic years, approximately 4.47 concussions occurred per 10,000 athlete exposures, resulting in around 10,560 concussions annually (Chandran et al., 2022; Edelstein et al., 2023). Women’s soccer ranked second among sports with the highest concussion rates (Zuckerman et al., 2015). In spite of differences in concussion exposure, NCAA concussion assessment protocols have tended to remain gender-agnostic, applying the same self-report diagnostic tools to male and female athletes alike (Beran & Scafide, 2022; Meehan et al., 2013; Kroshus et al., 2021).

Research consistently shows that male and female athletes differ in both the frequency and severity of reported concussion symptoms. Studies have indicated that female athletes tend to report a higher frequency of concussions, along with more severe symptoms, compared to their male counterparts (Granito, 2002; Broshek et al., 2005; Gessel et al., 2007; Iverson et al., 2021; Kieffer et al., 2021; Resch et al., 2017). These gender differences suggest that the current gender-neutral assessment methods may overlook key factors influencing symptom expression and recovery outcomes, potentially compromising the accuracy of concussion management (Covassin & Elbin, 2011; Brown et al., 2015; Cantu & Register-Mihalik, 2011). Furthermore, a systematic review of gender differences in SRC revealed that female athletes often experience longer recovery times, though the exact reasons for these differences remain uncertain (Edelstein et al., 2024). Such discrepancies may stem from biases in the diagnostic tools, which have historically been developed using predominantly male populations (Randolph et al., 2009; D’Lauro et al., 2022).

### The Sports Concussion Assessment Tool (SCAT)

The *Sports Concussion Assessment Tool (SCAT)* is a widely used assessment measure developed to systematically evaluate symptoms and cognitive functioning in athletes who may have experienced a concussion (Alla et al., 2009; Bretzin et al., 2022). Initially introduced in 2004, following *the 2nd International Symposium on Concussion in Sport*, organized held in Prague, Czech Republic, and subsequently updated (SCAT2 in 2008, SCAT3 in 2013, SCAT5 in 2017, and SCAT6 in 2023; The version number SCAT version 5 was chosen to align the version number with the *5th International Consensus Conference on Concussion in Sport*, held in Berlin, Germany, in 2016, meeting number and, as such, there is no SCAT4), the SCAT reflects advancements in concussion research and clinical feedback aimed at standardizing concussion assessment and improving its sensitivity. A depiction of the historical development of the SCAT components is presented in **Table 1**.

**Table 1:**
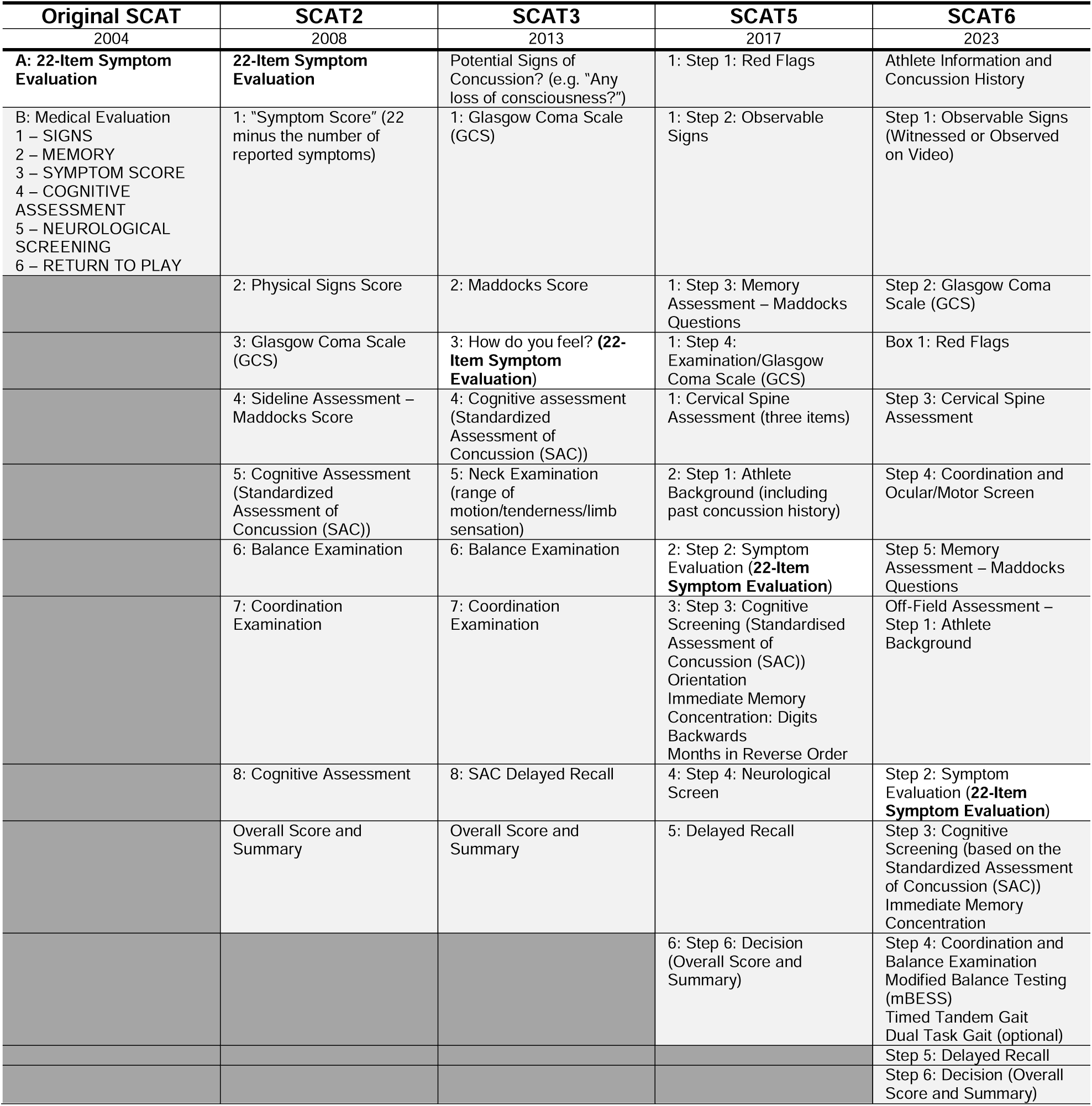
Versions of the Sports Concussion Assessment Tool (SCAT) While the 22-item self-report *Symptom Evaluation* component of the SCAT (shaded in white) has shifted its position in the list of assessments to be completed with each new version of the SCAT, the Symptom Evaluation component has remained as a consistent element of the instrument since its inception. As additional elements have been added to the SCAT, however, no sub-scaling of the Symptom Evaluation has implemented, nor has there been any inclusion of female-athlete specific components within the SCAT.

**Table 2:**
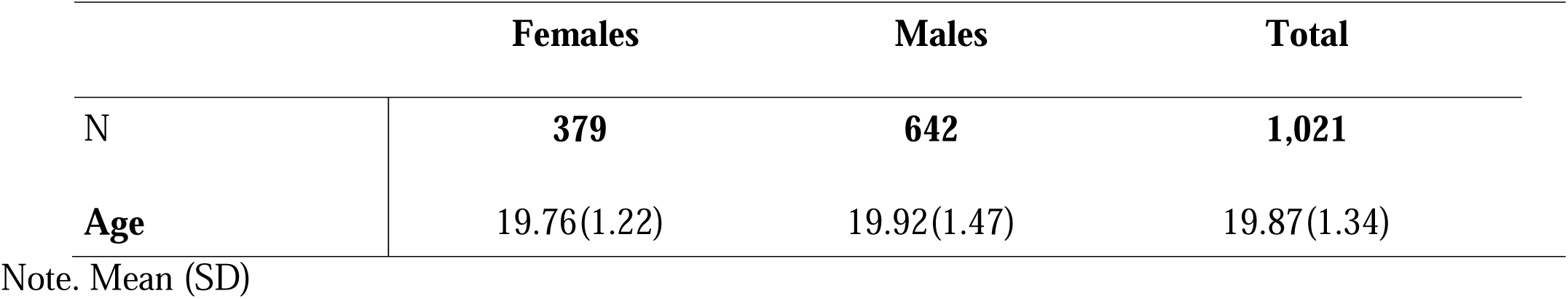
Demographics.

The SCAT incorporates both subjective symptom self-reporting and objective cognitive and physical assessments. The symptom evaluation component, which lists common concussion symptoms such as headaches, dizziness, nausea, and mental fog, requires athletes to self-rate the severity of each symptom. This self-reporting is meant to align with the recognition that subjective symptomology is an essential indicator of concussion severity and recovery. Alongside symptom evaluation, SCAT includes a cognitive assessment that tests immediate and delayed memory, concentration, and orientation, as well as balance testing through the modified Balance Error Scoring System (BESS). SCAT has been widely adopted in sports, particularly at professional and amateur levels, and is endorsed by major organizations such as the *International Olympic Committee (IOC)* and *World Rugby*.

The SCAT5 introduced several refinements over the SCAT3 to enhance usability and clinical accuracy. One notable change was the expansion of the symptom checklist to accommodate a broader range of neurological symptoms and increased sensitivity to symptom severity. This update was performed in response to research on the diverse ways concussions present, aiming to capture subtle changes that could be missed in earlier iterations. SCAT5 also offers updated guidance on interpreting symptom severity scores, including clearer thresholds to guide clinical decisions about when athletes should return to play. Additionally, it emphasizes cognitive and neurological examination components, enhancing sections on memory, concentration, and balance testing. For instance, the SCAT5 includes more detailed instructions for assessing balance using the modified Balance Error Scoring System (BESS), an essential metric for detecting vestibular and motor impairments post-concussion. Moreover, the SCAT5 provided updated guidelines on its utility across age groups, particularly recommending modifications for athletes under 13 with the Child SCAT5. Overall, changes in SCAT5 sought to provide a more robust framework, allowing clinicians to identify and manage concussions with greater confidence and precision compared to the SCAT3. The current, SCAT6 version (released in 2023), extends the set of neurological assessment domains still further (see below as well as in Table 1).

However, the self-report symptom rating portion of the SCAT has remained a consistent core feature across all iterations, from its initial version in 2004 to the SCAT6, despite numerous enhancements to other sections of the tool (Echemendia et al., 2017;“Sport Concussion Assessment Tool 6 (SCAT6),” 2023). This feature in the assessment reflects the central influence of subjective symptom reporting in concussion diagnosis and management, as athletes’ descriptions of their symptoms provide critical insights that objective or more clinical observation measures may not fully capture.

In the self-report portion of the SCAT, athletes rate the severity of various concussion symptoms—such as headache, nausea, dizziness, and cognitive fog—on a scale from 0 (none) to 6 (severe), creating a total symptom score. This scoring approach has not changed, even as additional assessment components, like more detailed cognitive testing, neurological assessment, and balance evaluations, have been added to enrich the SCAT’s comprehensiveness. Notably, throughout the history of the SCAT, there has never been any particular differentiation between the experience of female versus male athletes in response to the perceived symptoms of concussion and the assessment provides no sub-scaling nor differentiation between male and female symptom reporting.

### The NCAA-DoD Concussion Assessment, Research, and Education (CARE) Consortium

The NCAA-DoD *Concussion Assessment, Research, and Education (CARE) Consortium* represents a large-scale, multi-institutional research initiative focused on understanding SRC, primarily in college athletes and military cadets. Launched in 2014 as a collaboration between the NCAA and the U.S. Department of Defense (DoD), CARE aims to advance knowledge on concussion diagnosis, management, and recovery, as well as long-term health outcomes. This consortium integrates data from baseline assessments, injury evaluations, and post-injury follow-ups to study concussion trajectories comprehensively. Involving over 30,000 athletes across 30+ institutions, CARE represents one of the largest databases of concussion data globally. Its assessments encompass a wide range of modalities, including self-reported symptoms, neurocognitive testing, neuroimaging, and genetic analyses, allowing for a multidimensional understanding of concussion’s impact on brain health. The CARE Consortium collects pre-injury baseline data from athletes, enabling comparisons between pre-and post-concussion states and for tracking recovery progress in detail. This data-design has allowed for insights into gender, age, and sport-related differences in concussion risk and recovery, as well as the development of sophisticated predictive models for post-concussive outcomes. Through its broad dataset, CARE supports numerous research initiatives to develop evidence-based clinical guidelines, improve safety protocols, and identify biomarkers that could lead to more accurate diagnosis and personalized treatment for concussions, with impacts extending beyond sports medicine to military and civilian healthcare.

All data from the CARE Consortium are made openly available on the *Federal Inter-Agency Traumatic Brain Injury Resource (FITBIR)* data archive (fitbir.nih.gov). This includes clinical assessments, neuroimaging, balance test metrics, etc. The version of the SCAT used by the CARE Consortium for which data is available in the FITBIR archive was obtained using the SCAT3 version of the assessment, so while not reflective of the current state-of-the-art concerning the SCAT, it reflects the same core set of 22 self-report items as the more recent, SCAT6, assessment.

### Scoring the SCAT Self-Report Assessment

The SCAT6 incorporates several clinical and cognitive test components designed to provide a comprehensive assessment of concussion effects on an athlete. The clinical component begins with an “Immediate or On-Field Assessment” to identify any severe injury markers, termed “Red Flags,” such as neck pain or altered consciousness, which necessitate urgent medical attention. This is followed by an orientation and memory section, in which athletes answer basic questions regarding time, place, and recent events, aiding in the identification of disorientation or memory loss. The cognitive portion includes immediate memory recall, where athletes repeat a list of words presented to them, testing short-term memory, and a concentration test involving number sequencing and reverse recitation, which assesses focus and mental processing. Additionally, the delayed recall component tests retention by asking athletes to recall the initial list of words after a brief delay, providing insight into memory consistency over time. The SCAT6 also integrates a modified Balance Error Scoring System (BESS), which assesses postural stability by having the athlete balance in various stances while clinicians score any errors in posture or movement. Together, these tests evaluate an athlete’s cognitive functioning, memory, and balance—key areas frequently affected by concussion—helping clinicians make informed decisions about diagnosis, treatment, and readiness for return to play. Specifically, athletes complete a cognitive screening that includes orientation questions, immediate memory recall, concentration tasks (like “serial 7s” or months-in-reverse-order), and delayed recall; each scored separately. Physical testing, such as the modified Balance Error Scoring System (BESS), evaluates the athlete’s postural stability by measuring errors made during various stances.

Total scores on the SCAT do not yield a simple “pass/fail” outcome; instead, high scores indicate a generally more significant symptom burden and/or level of impairment. While no universal threshold score dictates whether an athlete is concussed, clinicians compare SCAT scores against baseline scores, if available, to detect changes and monitor recovery. However, the underlying basis of all versions of the SCAT checklist is the assumption that symptom severity can be captured by the *Total Symptom Score*, regardless of gender (Chin et al., 2016; Wilmoth et al., 2020). This overall number is then frequently employed to make clinical determinations on an athlete’s concussion severity and, ultimately, any clinical response. However, research by the CARE Consortium has suggested that the pooling of responses from across the range of SCAT items may not reflect the more subtle elements of head injuries in both male and female athletes (CARE Consortium Investigators et al., 2018; Nelson et al., 2018).

### Considering the Definition of Gender

The interchangeability of the terms “gender” and “sex” in SRC research further complicates the issue of considering the differential effects concerning a spectrum of gender identities. “Sex” refers to biological differences between males and females, while “gender” encompasses the social roles, behaviors, and identities associated with each (Heidari et al., 2016). Sports are commonly classified as “men’s” or “women’s” versions, based upon the biological interpretation. This distinction is nuanced but essential for understanding how social and cultural factors may influence symptom reporting and recovery. The SCAT also makes no attempt to capture differences which may be relevant to transgendered athletes. Beyond these issues, most modern concussion assessment tools, including the SCAT checklist, would ideally need to adequately consider how factors related to the gender of the athlete could skew symptom evaluation and recovery outcomes. In a broader sense, the consideration of SRC across the spectrum of perceived gender identities is beyond the scope, per se, of the present investigation, and the consideration of gender is limited to male and female labels, as reported by the CARE Consortium in the FITBIR archive.

### Examining the Structure of the SCAT and the Potential for Gender Differences

Under the SCAT assessment, female athletes are more likely to report a broader range of symptoms with greater severity than males (Guskiewicz et al., 2005, 2007; McCrea et al., 2003). Thus, these findings highlight the need to reexamine the psychometric properties of the SCAT checklist to ensure it accurately reflects the symptomatology of both genders (Randolph et al., 2009).

To address these gaps, the present investigation seeks to deconstruct the multidimensional nature of SRC symptom reporting, focusing on how gender may influence the perception and reporting of symptoms. By drawing on a robust dataset from the NCAA and DoD CARE Consortium, the study applies a suite of advanced multivariate statistical methods to assess the underlying dimensionality of concussion symptoms and to unravel the complexity of symptom reporting across genders. Exploratory Graph Analysis (EGA), Principal Component Analysis (PCA), Exploratory Factor Analysis (EFA), and Linear Discriminant Analysis (LDA) are employed to explore and clarify the dimensional structures that underlie the symptom clusters reported on the SCAT checklist, providing insights into how different symptoms co-vary and whether specific patterns emerge across male or female athletes.

As a complement to these methods, Differential Item Functioning (DIF) analysis and Rasch Modeling are used to rigorously investigate whether the SCAT checklist might disproportionately represent symptom severity scores in both male and female athletes, even when adjusting for differing individual trait level differences between genders. This layered approach aims to go beyond simple univariate symptom reporting or intensity comparisons and seeks to identify whether any gender-specificity exists in the underlying assessments themselves. Importantly, this study seeks to identify distinct clusters of concussion symptoms that more accurately reflect gender differences, helping to present a more nuanced, multidimensional framework for concussion assessment.

Given the SCAT’s historical consistency and comprehensive coverage of subjective concussion symptoms, a multivariate analysis of its self-report items is both timely and highly relevant, especially in light of the tool’s lack of adjustments or thresholds that account for the gender of the athlete. Since the SCAT self-report section has remained largely unchanged across iterations, this stability offers a unique opportunity for researchers to analyze symptom reporting trends over time and across diverse populations. Despite research showing that gender differences may influence concussion symptomatology and recovery trajectories, the SCAT does not differentiate scores or assessment criteria based on gender, potentially overlooking nuanced variations in symptom reporting between male and female athletes. A multivariate analysis could reveal patterns and dimensions within self-reported symptoms that vary by gender, identifying clusters or specific symptom profiles that might be more predictive of prolonged recovery in one group compared to the other. By examining the dimensionality of symptom reporting with statistical rigor, this approach could provide valuable insights that might improve individualized concussion management. Such an analysis could support the development of more tailored concussion guidelines, refining both diagnostic and recovery protocols to account for gender-related differences, ultimately enhancing the clinical utility of the SCAT for both male and female athletes.

## Methods

### Demographics

N=1,021 NCAA student-athletes (379 females and 642 males) completed the SCAT Version 3.0 (SCAT3) Symptom Severity Checklist within 48 hours post-concussion, which was obtained from the *Federal Interagency of Traumatic Brain Injury Research (FITBIR)* in collaboration with the NCAA and DoD CARE Consortium. As noted above, the checklist includes 22 symptoms, each assessed using a 7-point Likert scale ranging from 0-to-6. These symptoms are summed for a *Total Symptom Severity Score,* in which values may range from 0-to-132.

### Statistical Approaches

A systematic approach was utilized to evaluate the SCAT concussion assessment instrument’s underlying dimensionality. First, an exploratory graph analysis (EGA) was performed to illustrate the SCAT’s potential underlying multivariate structure (Golino & Epskamp, 2017). This was followed by a Principal Component Analysis (PCA) (Pearson, 1901), which formed the basis for a subsequent Exploratory Factor Analysis (EFA) (Spearman, 1904), which was used to determine which assessment items load the most onto latent symptom constructs (Rencher, 1992). A linear discriminant analysis (LDA) was also performed to determine the most discriminating SCAT items between males and females (Trendafilov & Gallo, 2021).

Lastly, the Masters (1982) Partial Credit Model (PCM) and Angoff’s (1981) Differential Item Functioning (DIF) analysis were conducted to confirm LDA results and provide greater specificity to gender-related symptoms on the SCAT symptom checklist most sensitive to differences between male and female athletes. PCA, EFA, and LDA analysis were also conducted through R version 4.2.2 (R Core Team 2022). In what follows, we describe the details involved in each of these steps:

### Exploratory Graph Analysis (EGA)

Utilizing R version 4.2.2 in conducting EGA, running the *EGAnet* package version 1.2.3 (Golino & Epskamp, 2017). These network-based models use nodes to represent random variables connected by edges, indicating the level of unique interaction between them rather than individuals in networks, aiding in determining the number of dimensions through cluster detection.

### Principle Component Analysis (PCA)

PCA was utilized for dimensionality reduction while preserving as much of the variability in the data as possible, deconstructing the item-wise correlation matrix, and transforming the original variables into a new set of linear combinations of the original variables. These new variables, called principal components (PCs), are orthogonal (independent of one another) and ordered, so the first few retain most of the variation in all the original variables. The eigenvalues associated with each PC were examined, and those greater than or exceeding unity (e.g., Kaiser’s Criterion) were taken as indicative of the SCAT assessment’s multivariate sub-space.

### Exploratory Factor Analysis (EFA)

Building from the PCA and assessing the number of putative factors, an EFA was done to determine the final number of factors and the subsequent standardized loadings for each assessment item, loadingonto each factor. Three and four-factor models with varimax rotation were compared by analyzing fit indices, computing the x2 fit statistic, RMSEA, the Comparative Fit Index (CFI), and the Tucker-Lewis Index (TLI) (Bakker et al., 2017).

To determine clusters of items and their corresponding factor, a cut-off score for the standardized factor loadings of 0.21 comes from the approach where the smallest acceptable absolute factor loading is determined as one over the square root of the number of items (Comrey & Lee, 1992). This is consistent with Huang (2018) concerning psychometric validation for ensuring robust factor interpretation and dimensionality.

### Linear Discriminant Analysis (LDA)

LDA finds the best linear combination of assessment items, which maximally separates two or more distributions relative to within-distribution variability (Rencher, 1992). Moreover, LDA is solving an eigenvector (*w*) problem to maximize group separation. To determine this separation, the coefficients and corresponding eigenvalues (λ) were used to determine directions along the axes, computing the identification of symptoms best separated by gender. The eigenvector coefficient equation comes from solving the eigenvalue equation (*A* - *λI*)*w* = 0, where A is a square matrix of the between-class and within-class scatter matrices (Xanthopoulos et al., 2013). The eigenvector coefficients are the elements of the eigenvector, and these coefficients can be found by solving the linear system derived from the matrix A (Trendafilov & Gallo, 2021). Furthermore, these coefficients define how the features combine to form the maximal gender separation between symptoms.

Finally, the statistical significance of this discriminant function was evaluated using Wilk’s Lambda (⋀) statistic and its approximate F-ratio test statistic. A low ⋀ value approaching 0 and a significant p-value indicates that the discriminant function explains a substantial portion of the variance between the groups (Klecka, 1980).

### Rasch Analysis

#### Rasch Partial Credit Model (PCM)

The PCM, a model within the family of Rasch measurement theories, was employed to analyze the response data (Masters,1982). This model is particularly suited for handling ordinal response categories typical of symptom severity scales, offering robust estimations of item difficulty parameters without assuming equal distances between each category. The PCM equation can be interpreted as follows: *r* is the current step, *x* is the current step, and *m* is the full set of categories. The numerator sums up to the current category, while the denominator is the sum of all the categories:

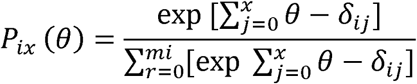

Delta parameters δ*ij* are specified per item; δ*ij* is the step difficulty or location where 2 categories intersect (category intersections). Item locations (*βi’s*) are typically obtained by taking an average of all the deltas (δ*ij*), the points along the latent trait continuum at which the likelihood of endorsing successive response categories increased (Embretson & Reise, 2000; De Ayala, 1989). The estimated thresholds for each symptom were used to identify the levels at which respondents were likely to move between response categories. Symptoms with disordered thresholds were marked for further review.

Before applying the PCM, the assumptions of unidimensionality, local independence, and monotonicity were tested. Unidimensionality was assessed through Exploratory Factor Analysis (EFA), ensuring that all symptom items measured a single latent trait (Embretson & Reise, 2000). PCM assumes that the item parameters (e.g., symptom difficulties and thresholds) are invariant across different genders. This assumption implies that the model should work equally well across genders. Lastly, evaluating the assumption of monotonicity ensures that as the underlying trait increases, the probability of endorsing higher response categories also increases (Masters, 1982; Tennant & Conaghan, 2007).

#### Differential Item Functioning (DIF)

DIF occurs when individuals from different groups (e.g., genders) respond differently to a symptom despite having similar underlying latent trait levels. In the context of concussion symptom reporting, DIF quantifies the extent to which male and female athletes may interpret or report symptoms differently. In such cases, one gender may be more likely to endorse a symptom at a higher severity level than the other, even though their actual level of concussion-related impairment may be the same (Bollmann et al., 2018; Masters, 1982). While measurement invariance across groups and time is desirable, cases in which symptoms are endorsed more severely for one gender versus another indicate lack of support for it.. Ensuring measurement invariance is essential to ensure that the assessment accurately reflects an equivalent probability of endorsement for items for all individuals, regardless of gender (Embretson & Reise, 2000). The following equation further computes the overall DIF measure or the difficulty for each gender and corresponding item

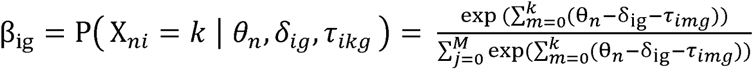

The DIF equation for the PCM involves several key variables (Masters, 1982). To compute the DIF measure for the corresponding gender and item β_ig_, the θ_n_ takes the ability level of person *n*, while δ*_ig_* is the difficulty of the item within each gender, *g*. The τ_img_ are step thresholds that define the boundaries between response categories.

DIF contrast quantifies the difference in item difficulty between groups (e.g., genders) for a specific symptom. The size and direction of the contrast indicate whether and to what extent males and females tend to report a higher severity (Dorans & Holland, 1992). The equation below shows how the DIF contrast was computed, which takes the difference between the item difficulty for the prechosen reference group (females) and the focal group (males). This contrast value helps identify DIF direction and assess whether different genders respond differently to the same item after controlling for trait level (Bollman et al., 2018). This is found using the following Masters (1982) equation

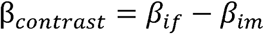

Where *i*= item, *f*= female, and *m*= male. A negative contrast value of *β_if_* − *β_im_* suggests that females tend to more frequently report the symptom as relatively more severe than males at the same level of concussion severity (Angoff, 1981). In other words, for the same overall concussion effects, females are more likely to endorse higher ratings for that symptom than males. Contrasts in the opposite direction, or a positive contrast, suggest that males tend to more frequently report the symptoms as relatively more severe than females.

To further determine significant symptoms that display DIF, Mantel-Haenszel probability statistics were used to determine whether an item exhibits uniform DIF between two observed groups, that is, whether an item is more frequently endorsed by one gender relative to the other, considering the latent trait. To avoid alpha inflation and Type I errors stemming from multiple comparisons, Benjamini & Hochberg (B-H) *post-hoc* tests (Benjamini & Hochberg, 1995) were conducted, as testing item DIF for many items poses an increased risk of Type I errors due to multiple tests with α < .05. Therefore, B-H correction was appropriate to control the false discovery rate (FDR) associated with the multiple comparisons (Buchardt et al., 2023).

## Results

### Demographics

As seen in **Table 3**, this analysis revealed a statistically significant difference in the mean Total Symptom Severity Scores between females (M = 30.06, SD = 20.88) and males (M = 24.71, SD = 21.18), t (765.06) = 3.85, p < .001.

**Table 3.**
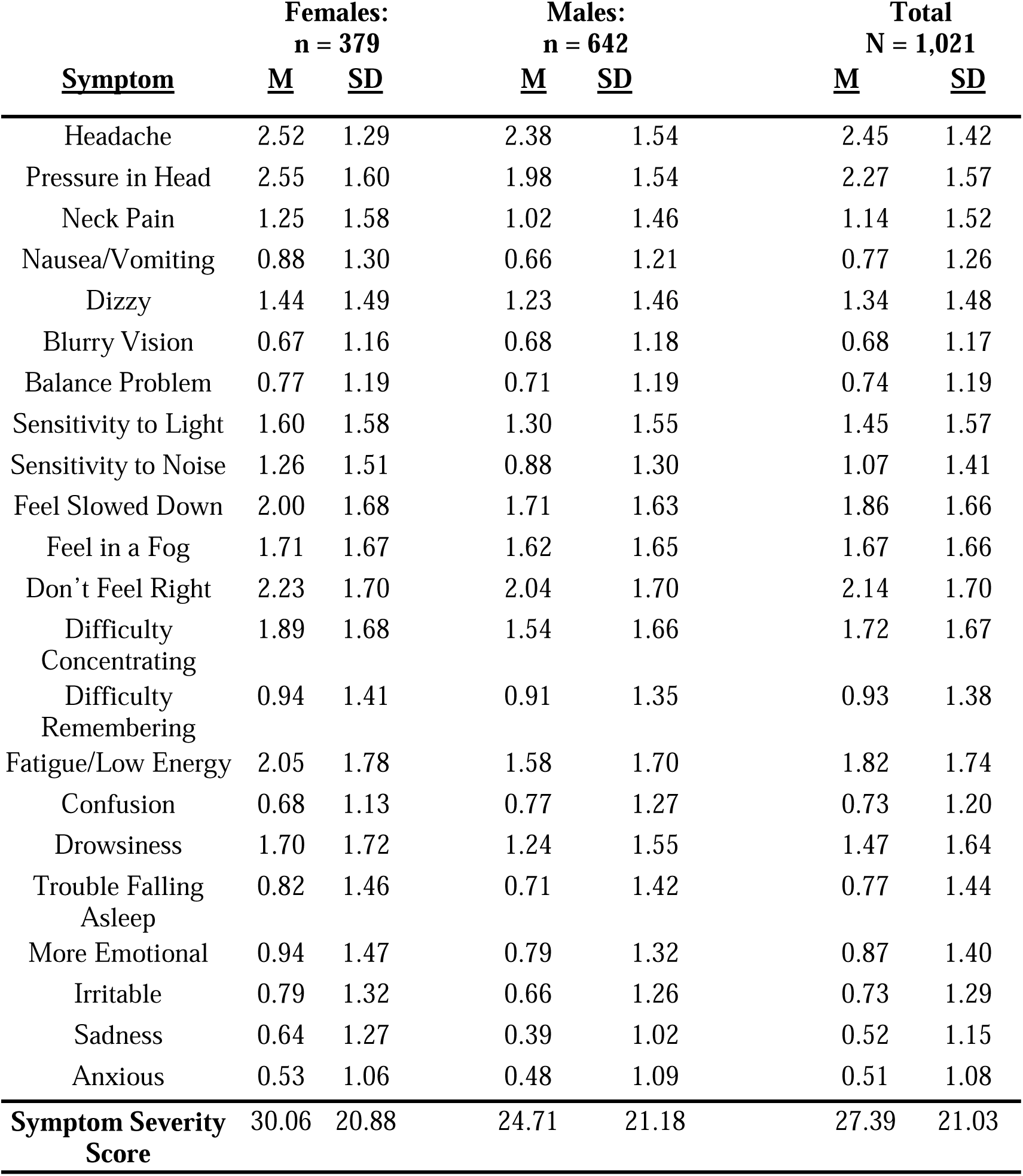
Item Descriptive Statistics.

### Dimensionality of the SCAT Self-Report Items

#### Exploratory Graph Analysis

The EGA network visualization and corresponding network loadings represent different symptoms or states grouped into clusters based on their underlying correlations (**Figure 1**). The analysis identified five clusters. However, one factor contained only three items, and two had relatively low loadings, minimizing the variance. This was taken as justification for further analysis using PCA and EFA to achieve a more parsimonious factor structure, reflecting symptom reporting sub-spaces in the SCAT.

**Figure 1:**
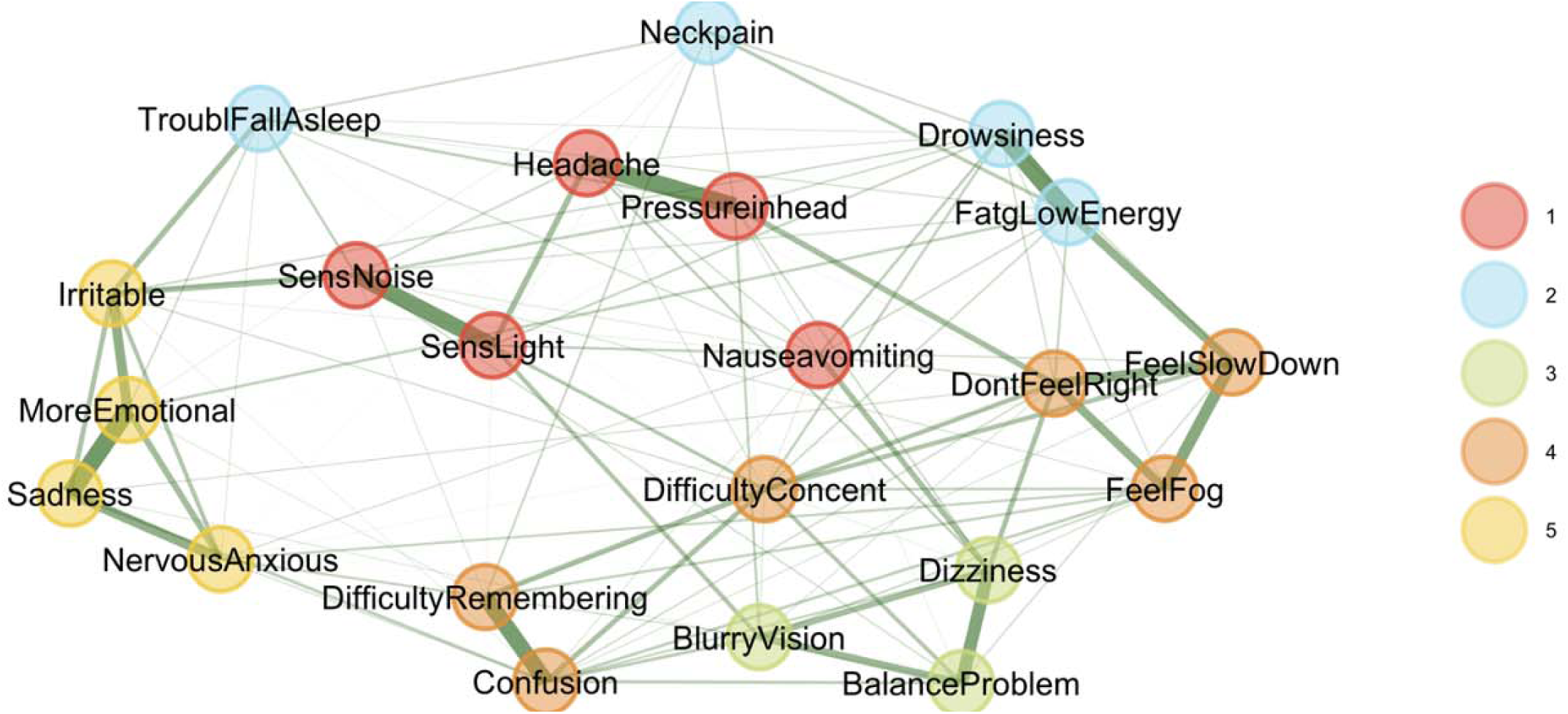
Exploratory Graph Analysis. This graph illustrates the SCAT3 Symptom Checklist, revealing three correlated fact rs. Each node represents a symptom, with lines indicating relationships between symptoms based on the stages the instrument intended to measure.

#### Principal Components Analysis

PCA identified four significant components, as rendered through a scree plot (**Figure 2**) which were 9.82, 1.63, 1.20, and 0.998, respectively; the fourth component’s eigenvalue nearly meets the Kaiser criterion, suggesting potential additional information. The fourth dimension explains 4.53% of the variance, leading to a higher cumulative explanation of 62.44% in the four-factor model. Including the additional fourth factor may result in a more comprehensive representation of the dataset, ensuring that subtler yet important patterns are accounted for.

**Figure 2.**
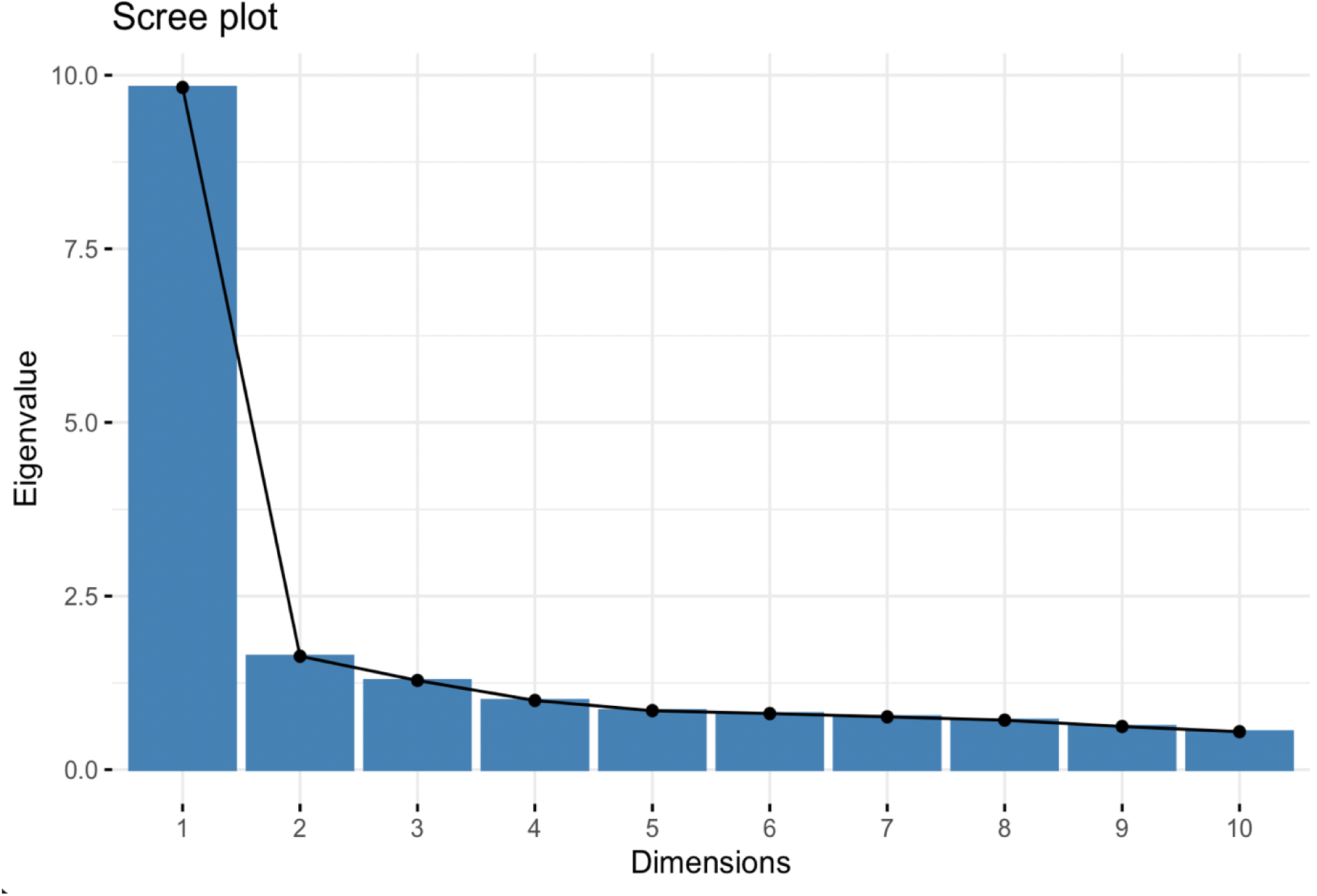
PCA Screeplot. The screeplot displays eigenvalues (9.82, 1.63, 1.20, 0.998) against the dimensions, helping to identify the four factors to retain in exploratory factor analysis.

#### Exploratory Factor Analysis (EFA)

Two models, a three-factor and a four-factor model, were compared to identify the most suitable number of factors. The four-factor model demonstrated a significantly better fit, as the three-factor model (*x*^2^(168) = 1,527.04, CFI= .89, TLI=.85, RMSEA= .09) demonstrated lower fit indices compared to the four-factor model to (*x^2^*(149) =957.23, CFI=.94, TLI=.90, RMSEA=.07). The statistical improvements in fit support the inclusion of a fourth factor, as it better captures the multivariate subspace of the data. In turn, if the inclusion of an additional factor aligns well with known constructs, it is often best to include it. In concert with these fit values, the PCA results indicating a fourth eigenvalue close to unity suggest a four-factor model will result in a more parsimonious model.

#### Latent Factor Names

Based on the factor analysis shown in **Table 4**, the latent factors labeled as Neurocognitive, Neurophysiological, Neurosensory, and Neuropsychiatric were selected to encompass various dimensions of post-concussion symptoms. Each factor aggregates specific symptoms based on their underlying relationships and shared characteristics, providing a comprehensive understanding of the multifaceted impacts of concussions. The prefix “Neuro-” in each factor name effectively underscores the neurological basis of the symptoms associated with post-concussion syndrome:

1. *Neurocognitive*: Cognitive impairments, such as difficulty concentrating, memory problems, and mental fog, typically manifest from neurological disruption following a concussion. Labeling this factor as “neurocognitive” highlights the brain-based origin of these dysfunctions.
2. *Neurophysiological*: This factor includes symptoms that, while physical (e.g., headaches, sensitivity to light and noise), are a direct result of neurological damage resulting from concussion. This emphasizes that these symptoms are linked to neurological processes related to SRC, not just physical ailments involving other parts of the body.
3. *Neurosensory*: Symptoms grouped under this factor (e.g., blurry vision, balance problems, dizziness) are sensory-related and tied to the sensory pathways in the brain affected by the concussion. “Neurosensory” underscores the neurological origin of these sensory disturbances.
4. *Neuropsychiatric*: Emotional and behavioral symptoms (e.g., irritability, sadness, anxiety) often have neurological underpinnings. Labeling them as “neuropsychiatric” acknowledges that these symptoms are psychiatric potentially resulting from their SRC.

**Table 4.**
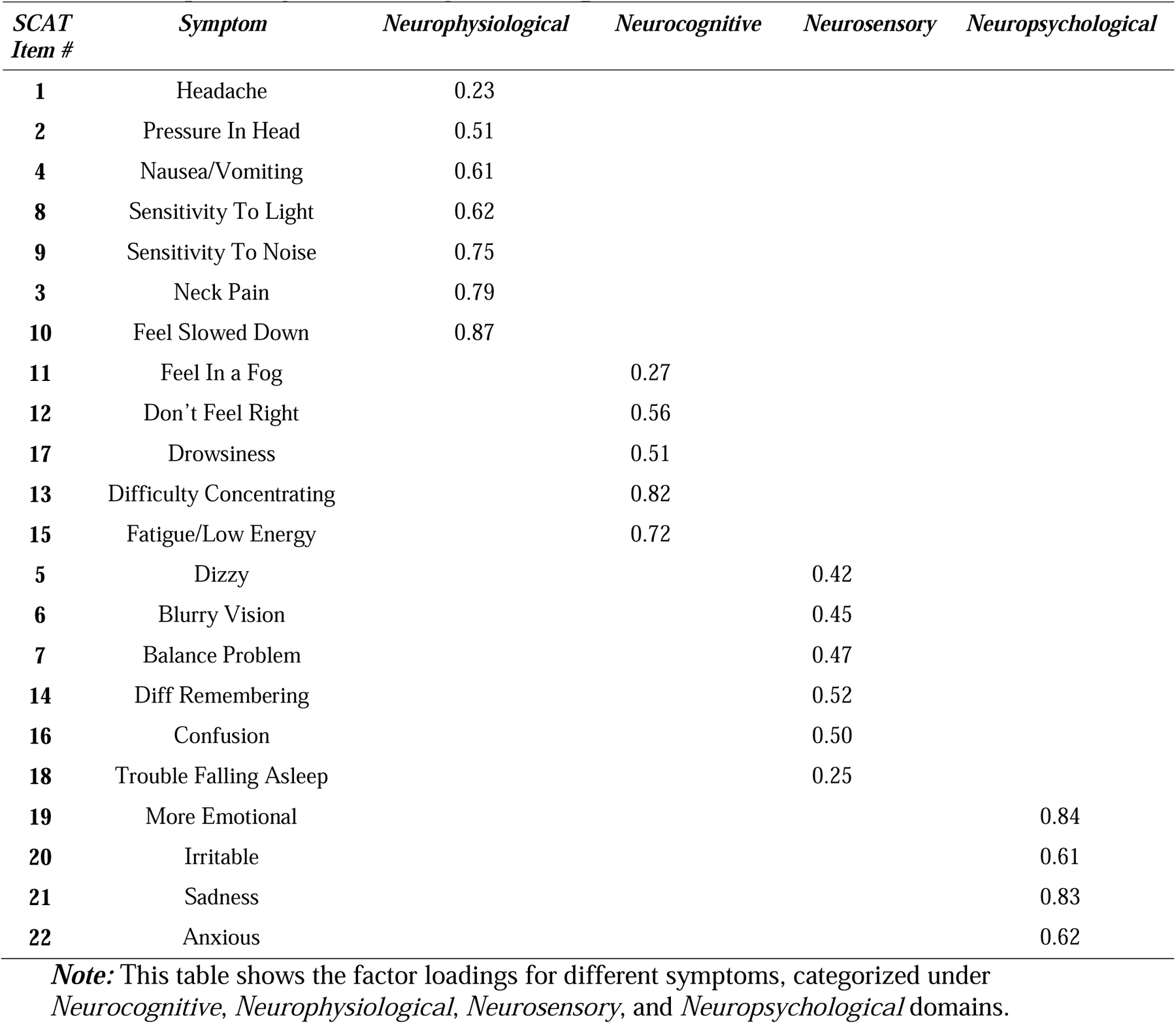
Exploratory Factor Analysis Loadings with Latent Variables.

Thus, each factor aggregates specific symptoms based on their underlying relationships and shared characteristics with respect to self-reported ratings. Importantly, males and females tend to load onto these factors differently which prompts further examination into those items which may be driving these differences.

#### Linear Discriminant Analysis (LDA)

LDA was conducted to identify items which, in a weighted linear combination, maximized differences in how groups responded and to identify potential item biases in advance of conducting a more item-specific DIF analysis (see below). The LDA revealed significant differences in symptom reporting between males and females (Wilk’s Λ = 0.82, χ² (22) = 130.56, p < .001), with an accuracy of 91% in distinguishing between the two groups. Thus, the differences between the groups are statistically significant and that the discriminant function is capturing meaningful distinctions between the genders, even though the effect size might be modest. Therefore, symptoms such as emotional distress, pressure in the head, sensitivity to noise, drowsiness, and sadness contributed most to these differences, with females reporting having these symptoms more frequently (**Table 5**).

**Table 5.**
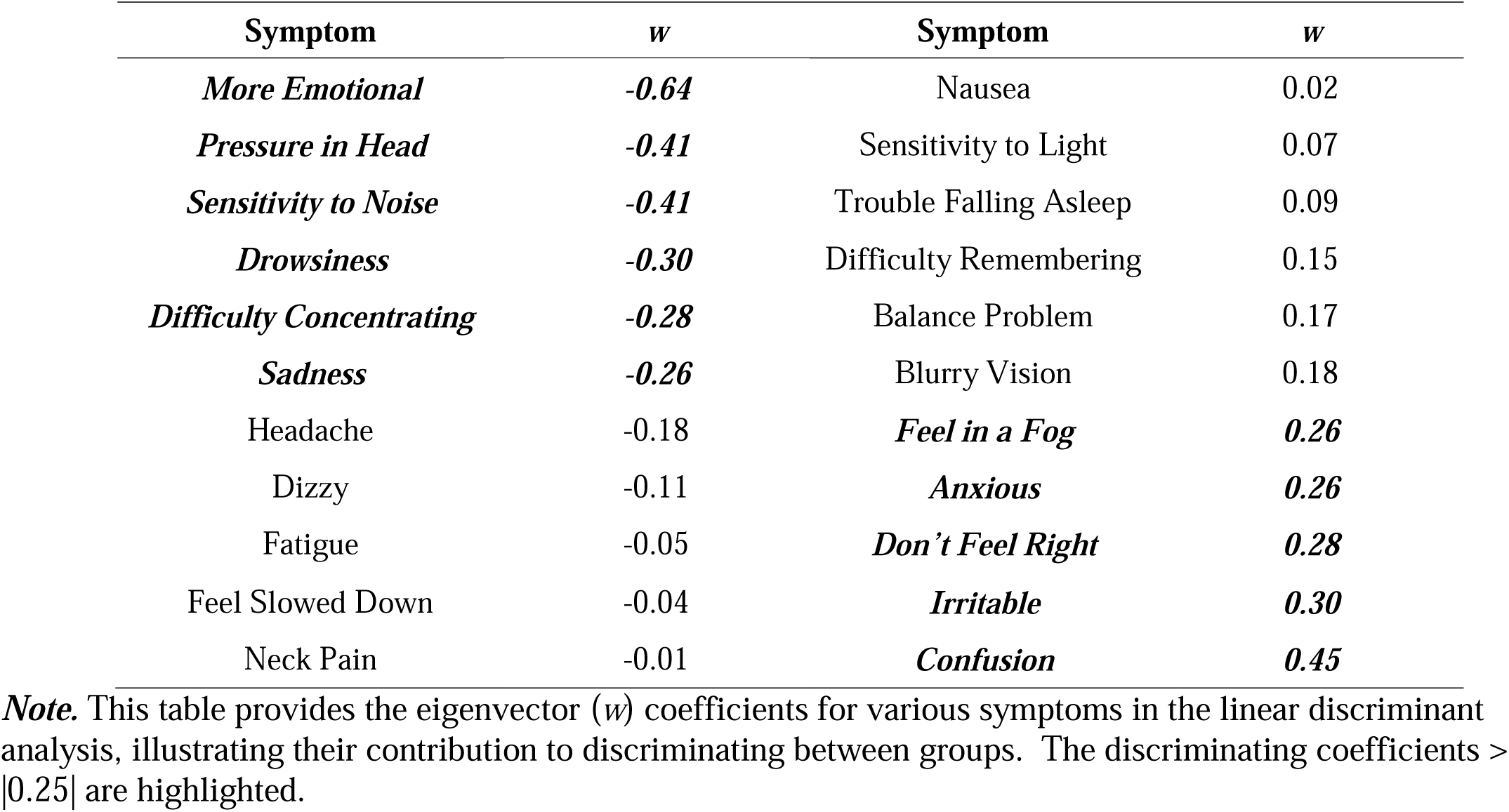
Linear Discriminant Analysis Eigenvector Coefficients.

This density plot in **Figure 3** of the LDA-derived distributions illustrates the separation of optimized symptom scores between male and female NCAA athletes. The overlap indicates some shared symptom presentation across genders. Still, the shift in the peak density for males compared to females suggests that females report specific symptoms at a slightly higher discriminant score, reflecting potential differences in symptom severity or reporting behavior between the two groups. Thus, the discriminant function holds practical importance in differentiating males from females in terms of symptom reporting.

**Figure 3:**
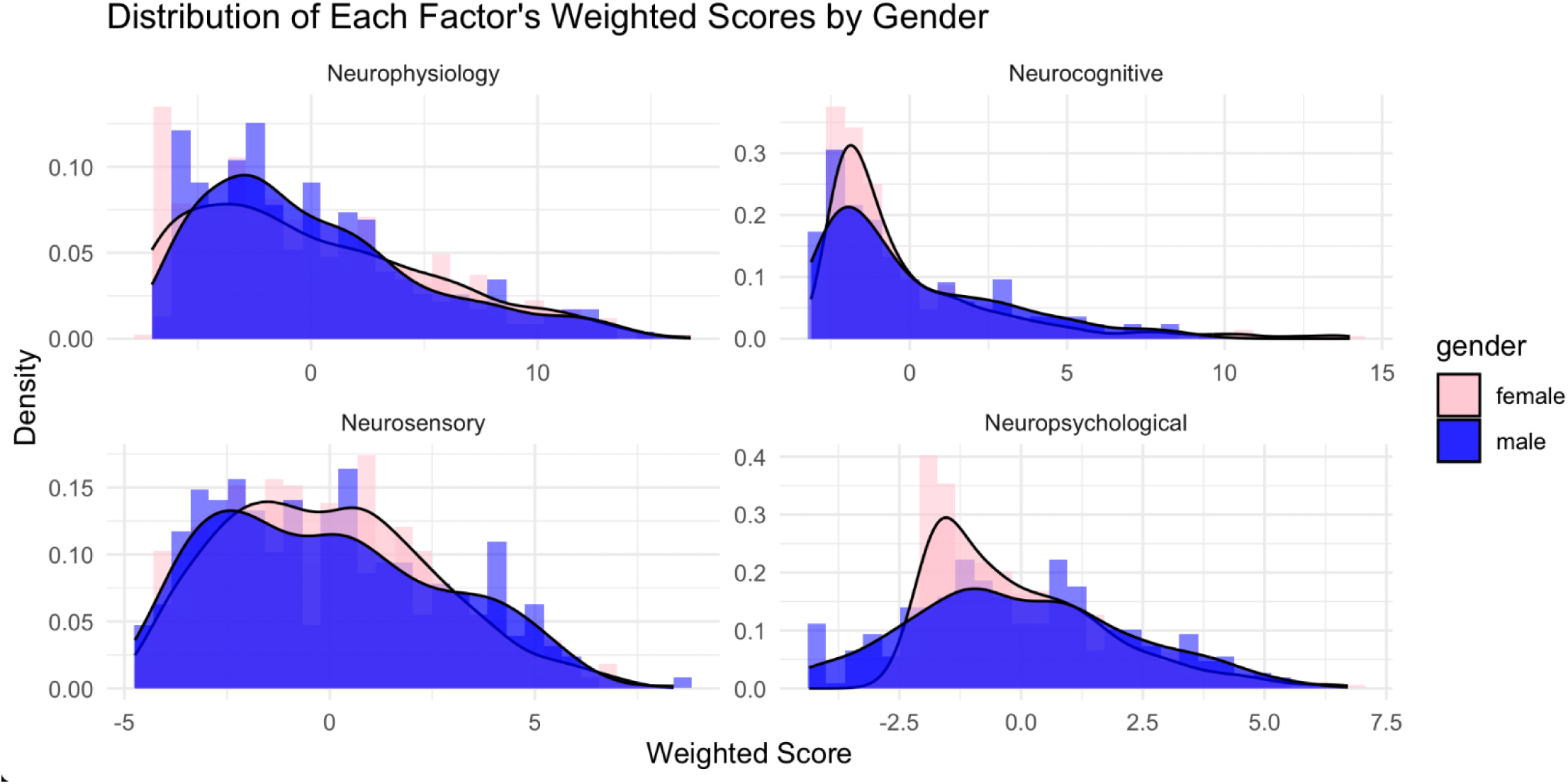
Factor Score Distributions by Factor by Gender. Factor scores were computed on each of the, so-named, *Neurophysiology*, *Neurocognitive*, *Neurosensory*, and *Neuropsychological* factors using the extracted weightings as presented in Table 3. (blue) and females (light red) are distinctly shown.

**Figure 4.**
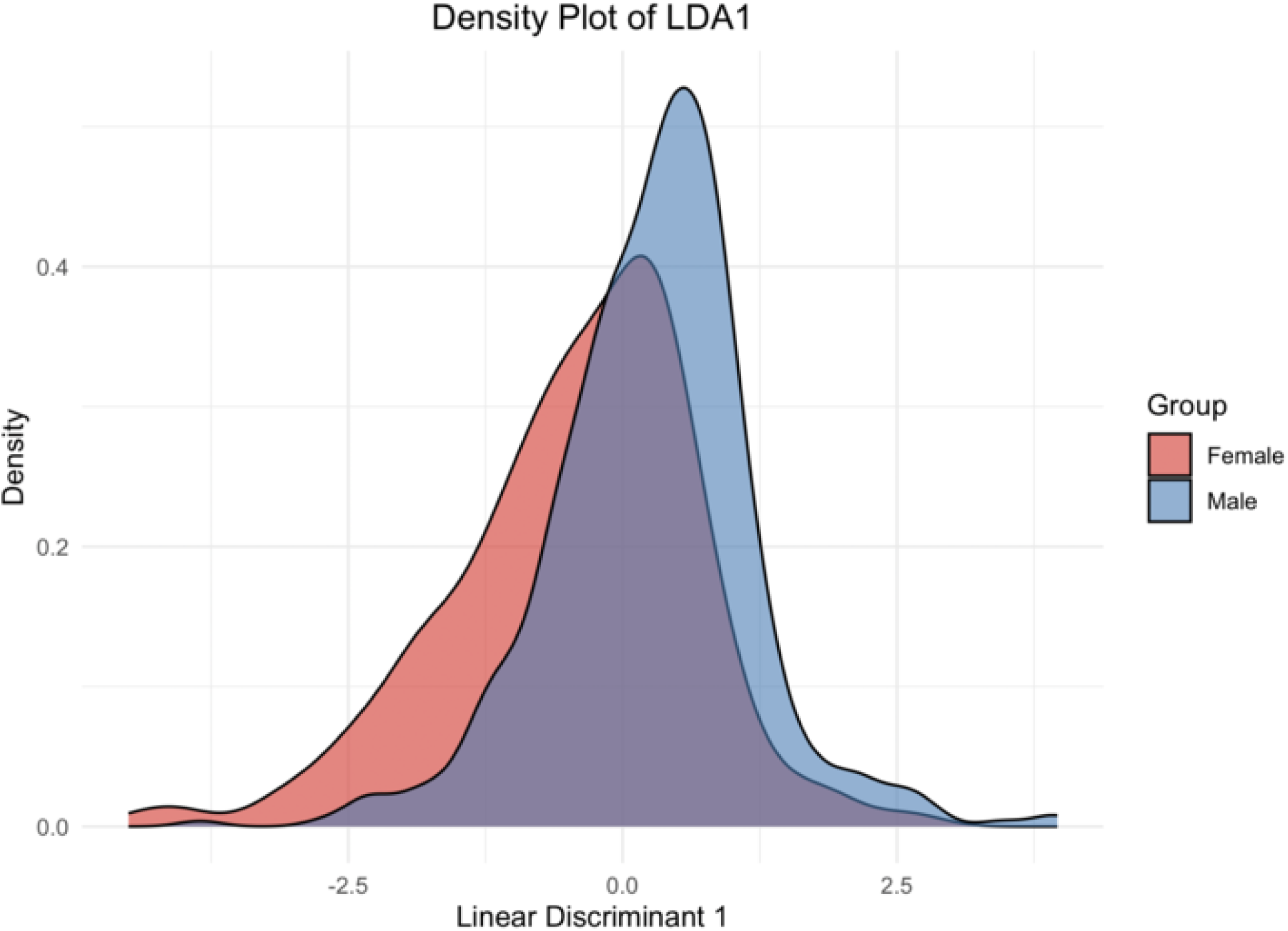
Linear Discriminant Density Plot. The graph shows how the linear discriminant function distinguishes between the male and female groups based on the underlying symptoms or features included in the model. The **peaks** of the curves indicate the most common LDA1 scores for each gender, while the **spread** of the curves reflects the variability within each group.

#### Rasch Partial Credit Model (PCM)

This Rasch model analysis utilizing the Partial Credit Model (PCM) was conducted through WINSTEPS Version 5.8.5 (Linacre, 2023) to examine the item thresholds for various symptoms related to a specified condition. The analysis delineated at which points along the latent trait continuum individuals were more likely to endorse successive response categories for each symptom, thus providing insight into the differential sensitivity of items as the underlying condition intensifies (De Ayala 1989; Embretson & Reise, 2000).

The results in **Table 6** indicated that the “Headache” has step difficulties ranging from -3.89 to 1.98, suggesting a wide range where this symptom progressively becomes more likely to be reported as the latent trait level increases from very low to high. For “Pressure in Head,” step difficulties spanned from -2.8 to 2.26, demonstrating that this symptom is relevant across a broad spectrum of the latent trait severity, with both genders starting to report this symptom at moderately low levels of the underlying trait. However, a few items displayed non-monotonic step difficulties patterns, where athletes report specific symptoms more efficiently at low severity levels but underreport them at moderate or high severity levels. Symptoms like “Nausea/Vomiting,” “Irritability,” “Sadness,” “Neck Pain,” and “Balance Problems,” respectively, suggested varied levels of the trait.

**Table 6.**
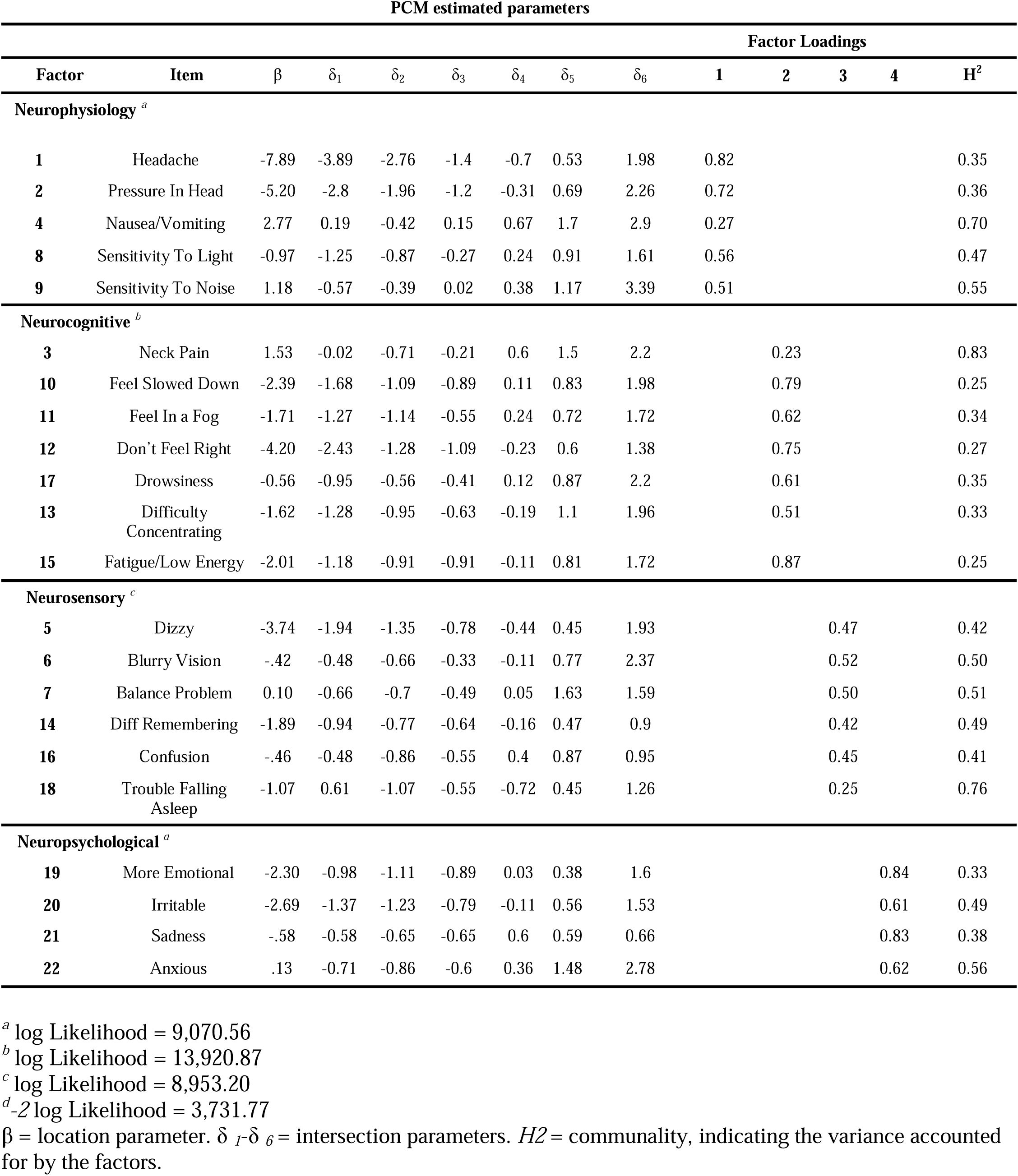
Partial Credit Model Estimated Parameters and Factor Loadings.

In examining monotonicity, most items indicated that as severity levels increase, the frequency of reporting symptoms as severe decreases. However, several symptoms display varying step difficulties, meaning reporting symptoms as a 1-2 or 5-6 symptoms may be more easily endorsed; however, reporting symptoms as moderate may be more difficult. For example, the symptom “Nausea/Vomiting” shows distorted thresholds as reporting symptoms as a 1 (δ_1_=.19) or a 3 (δ_1_=.15) and was more difficult to endorse than symptoms as 2 (δ_1_=.-42). As a result, certain symptoms may exhibit nonlinear characteristics. This means the relationship between these symptoms and their underlying causes does not follow a straightforward, predictable pattern.

#### Differential Item Functioning (DIF)

The results highlight significant gender differences in the reporting of concussion symptoms, with notable findings in several key symptom dimensions. DIF analysis reveals that specific symptom dimensions—such as neuropsychological, neurosensory, and neurocognitive—exhibit the most substantial gender-based differences in symptom reporting.

**Table 7** shows that nine symptoms in three dimensions exhibited significant DIF. Symptoms such as “Feel in a Fog,” “Don’t Feel Right,” “Fatigue,” “Dizzy,” “Balance Problem,” and “Anxious” exhibit positive levels of DIF, with female athletes consistently having reported lower symptom severity These findings support the PCM findings, as symptoms such as “Feeling in a Fog,” “Fatigue,” and “Balance Problem” indicating that males report higher severity more frequently, potentially due to underreporting at lower levels. In contrast, other symptoms such as “Difficulty Concentrating,” “More Emotional,” and “Confusion” demonstrate DIF in the opposite direction, indicating that male athletes are significantly less likely to endorse these symptoms at higher severity levels compared to females.

**Table 7.**
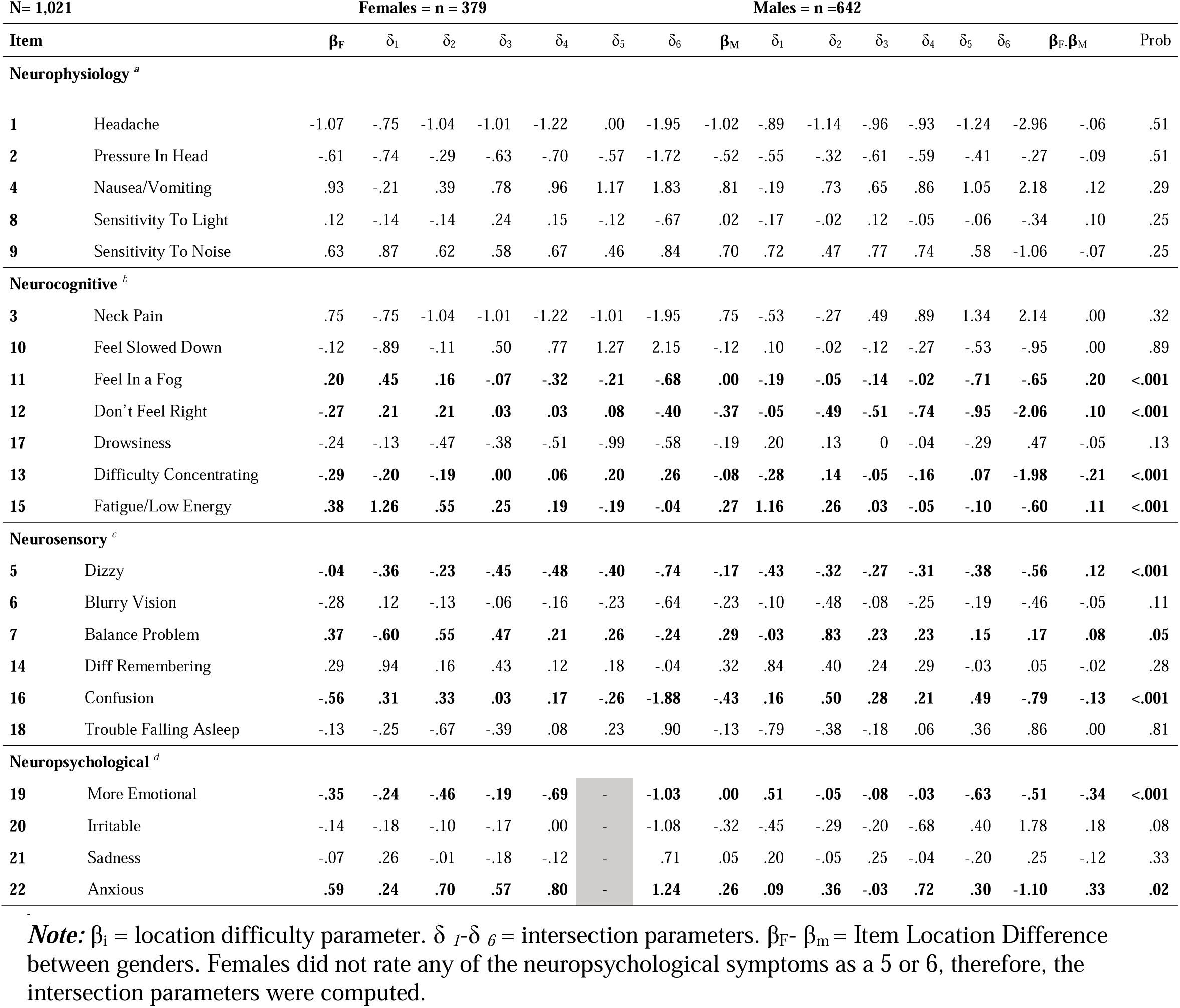
Differential Item Functioning Analysis Parameters.

These results, in addition to those of the LDA, provide robust evidence for gender differences in concussion symptom reporting, particularly in how male and female athletes respond to varying levels of symptom severity. Moreover, males’ symptom profile post-concussion aligns within cognitive and sensory domains, which might not affect their overall perception of symptom burden as much as the emotional and physical symptoms reported by females. Furthermore, the inflation in total symptom scores for females could be indicative of a reporting pattern where more diffuse symptoms contribute to a perception of greater severity, potentially leading to an overestimation of symptom burden.

## Discussion

The SCAT assessment has, through several revisions, proven to be a valuable, structured tool for evaluating SRC, offering clinicians a comprehensive framework to assess various symptoms and cognitive impairments associated with head injuries. Its multimodal design—incorporating symptom self-reporting, cognitive tests, and physical balance assessments—enables it to capture a wide spectrum of concussion-related effects, making it adaptable to on-field evaluations and post-injury follow-ups. The SCAT is particularly useful in providing both a baseline and post-injury comparison for athletes, supporting clinicians in detecting subtle cognitive and functional changes. By including items such as memory recall, concentration tasks, and balance assessment, the SCAT allows for a nuanced view of concussion impact, essential for understanding an athlete’s functional status and planning their safe return to play. This tool has become a standard in sports settings and continues to serve as a consistent, reliable resource in concussion assessment across different age groups and levels of athletic competition.

The SCAT’s utility is further supported by its structured and reproducible approach to concussion assessment, which promotes clinical consistency and reliability in a high-stakes environment where rapid, accurate decision-making is critical. By incorporating immediate indicators of severe injury, such as the “Red Flags” section, the SCAT aids in quickly identifying athletes who may require urgent medical care. Meanwhile, its core symptom and cognitive evaluation sections align with widely researched concussion symptoms, providing evidence-based guidance for diagnosis and management. However, while the SCAT’s general utility and standardized format offer clear advantages, its lack of tailored scoring based on factors such as gender, age, or sport type leaves room for further enhancement. The addition of these considerations could improve the SCAT’s sensitivity to variations in symptom presentation, thereby increasing its value as a diagnostic and monitoring tool and contributing to the continued development of more personalized concussion care in sports medicine.

However, the analyses performed here on the SCAT’s self-reporting portion reveal that this portion of the SCAT – the most consistent component over its history - encompasses a more nuanced set of symptom sub-scales within its broader collection of symptom ratings, highlighting complexities not captured by its single aggregated score. Certain symptom clusters—such as those related to mood, migraine-like pain, and cognitive fogginess—tend to emerge as distinct factors within the overall self-reported symptomatology, suggesting that a multivariate understanding could better capture the multidimensional experience of concussion. Additionally, male and female athletes often report differing symptom severities on specific items, with female athletes generally rating symptoms like headache, nausea, and emotional sensitivity more intensely than their male counterparts. These differences point to a need for gender-specific symptom profiling within the SCAT’s self-report section to capture the variations in how concussions manifest and are experienced across athletes.

Recognizing and addressing these sub-scales and gender-related response differences could make the SCAT a more finely tuned tool for assessing and monitoring concussion, leading to more individualized and effective clinical management.

Multivariate analyses revealed the presence of four distinct symptom clusters, reinforcing the argument for a multidimensional approach to concussion symptom assessment. Prior studies support that an effective concussion care protocol must acknowledge the neurocognitive, neurophysiological, neurosensory, and neuropsychiatric dimensions of symptoms (Kroshus et al., 2021; Nelson et al., 2018). The unidimensional model employed by SCAT3 is, by the analyses presented here, likely insufficient to capture the intricate interplay among these dimensions, potentially leading to misdiagnoses or mismanagement in athletic settings.

### SCAT Self-Report Sub-Scales

In our view, the SCAT assessment can be better understood through several extracted sub-scales which reflect distinct domains of concussion-related impairments. As noted above, these sub-scales not only capture unique components of variation in SRC self-report but items on which female athletes may respond differently. In the list below, we summarize these sub-scales and which items may combine to highlight gender-differences in SRC reporting (as determined by LDA presented in italics) or which show item-specific variations (indicated by DIF in bold):

#### Neurocognitive

This collection of items encompasses cognitive symptoms such as difficulty concentrating, memory issues, and mental fog, all of which are indicative of neurological disruption following a concussion. This includes the SCAT self-report items Headache, *Pressure In Head*, Nausea/Vomiting, Sensitivity To Light, *Sensitivity To Noise*, Neck Pain, and Feel Slowed Down.

#### Neurophysiological

This factor includes physical symptoms that, while experienced in the body (like headaches, sensitivity to light, and noise sensitivity), stem from neurological damage directly associated with concussion. Items on this sub-scale include ***Feel In a Fog***, ***Don’t Feel Right*, *Drowsiness***, ***Difficulty Concentrating***, and **Fatigue/Low Energy**.

#### Neurosensory

Symptoms in this sub-scale include blurry vision, balance problems, and dizziness, which are sensory in nature but linked to the brain’s sensory pathways impacted by the concussion. The items **Dizzy**, Blurry Vision, **Balance Problem**, Difficulty Remembering, ***Confusion***, and Trouble Falling Asleep comprise this sub-scale.

#### Neuropsychiatric

This sub-scale captures emotional and behavioral symptoms, such as irritability, sadness, and anxiety, which are often influenced by concussion-induced changes in brain function. Items included on this sub-scale include ***More Emotional***, ***Irritable***, *Sadness*, and ***Anxious***.

These sub-scales provide a structured means for interpreting the range of symptoms reported in the SCAT, allowing for a more granular understanding of the diverse effects concussions have across cognitive, physical, sensory, and emotional domains, and, notably, those items where females may respond differently to males.

### Gender Differences in SRC Self-Reporting

The present investigation highlights significant gender disparities in post-concussion symptom reporting among a large sample of NCAA student-athletes obtained, underscoring the limitations of the SCAT3 Symptom Severity Checklist’s traditionally scored unidimensional structure. Female athletes demonstrated a higher overall symptom severity, particularly within the emotional and sensory domains, suggesting an inherent bias in symptom assessment that warrants further clinical attention. These findings are crucial to understanding the implications of personalized concussion management strategies, especially given the historical tendency to underestimate the severity of symptoms in male athletes.

Additionally, it has been suggested that gender differences in symptom reporting may stem from cultural influences, with male athletes often underreporting symptoms due to societal pressures to exhibit “toughness” in competitive environments (Master et al., 2021). Such behavioral discrepancies can compromise the accuracy of symptom assessment and contribute to inflated severity scores for female athletes within the critical period following injury. This calls for a reevaluation of current assessment tools, as failing to account for these gender-based differences may result in male athletes being inaccurately perceived as less symptomatic, thereby jeopardizing their health and recovery trajectory.

The evidence emphasizes the need to modify existing concussion assessment methods, particularly the SCAT3. Clinicians must adopt a nuanced understanding of symptom reporting that integrates gender-specific considerations to enhance the accuracy of diagnoses, inform appropriate management protocols, and ultimately improve outcomes for all athletes.

### Implications for Female Athletes

The results of this study have several specific implications for the diagnosis, management, and treatment of female athletes with SRC. Firstly, the findings from this examination underscore the need for gender-sensitive approaches to concussion assessment. The SCAT self-report items, while widely used, may not be sufficient to capture the full spectrum of symptoms experienced by female athletes when used in its unidimensional form. Clinicians may need to account for the higher likelihood of emotional and sensory symptoms in females, which could contribute to a higher total symptom score but may not necessarily reflect more severe neurological impairment. Future revisions of the SCAT self-report questions and other concussion assessment tools should consider including gender-specific norms or symptom weightings to improve diagnostic accuracy and provide a more comprehensive assessment with respect to concussion symptoms experienced by women (McCrory et al., 2017).

Secondly, the results suggest that female athletes may require more individualized post-concussion considerations. The presence of emotional symptoms, such as anxiety and sadness, reported more frequently by female athletes, emphasizes the importance of providing comprehensive mental health support as part of concussion recovery. Additional psychological counseling, monitoring for depression and anxiety, and ensuring that emotional symptoms likely need particular attention and best not overlooked during clinical evaluations in female athletes (Wallace et al., 2017).

Lastly, little is known about the manifestation of longer-range neurodegenerative diseases in female athletes. One of the few studies found that female athletes who reported increased symptom severity within the first 48 hours following sports concussion (O’Connor et al., 2017), found that this could be an early indicator of more significant neuroinflammation or diffuse axonal injury, both of which are important precursors for long-term complications like chronic traumatic encephalopathy (CTE).

While symptom profiles provide important subjective insights into the athlete’s condition, they are best used with neuroimaging techniques to provide a more comprehensive brain injury assessment. Neuroimaging, particularly advanced techniques like functional MRI (fMRI), diffusion tensor imaging (DTI), and positron emission tomography (PET), can detect subtle changes in brain structure and function that might not be immediately evident from symptoms alone (Edelstein et al., 2024). For example, Asken et al. (2018) suggested combining symptom data with neuroimaging could enhance early identification of pathophysiological changes that may lead to CTE. By integrating symptom profiles—such as those identifying the neurocognitive, neurophysiological, neurosensory, and neuropsychiatric dimensions—with quantitative neuroimaging biomarkers, clinicians can create more accurate timelines for the onset and progression of CTE.

### Limitations of the Present Investigation

This investigation was conducted on NCAA athletes, and it is important to note that these results cannot be generalized to youth, high school, or professional sports. Most of the participants in the CARE Consortium sample attended academic institutions with well-funded athletic programs, which may reflect a greater attention to concussion symptoms, better quality of treatment, and more formalized programs for concussion management. Therefore, it is unclear if such results would be the same or similar in participants drawn from smaller athletic programs, historically Black colleges and universities (HBCUs), or other ethnically unique programs. Consequently, it would be advantageous to conduct further studies on more comprehensive, national samples including different age groups, contact vs. non-contact, youth, collegiate, and professional sports, as well as socioeconomic backgrounds to better understand and generalize the properties of SCAT as they pertain to both male and female athletes.

Indeed, this examination could not consider any psychological or social factors impeding symptom reporting. It will be important for future research to include variables that reflect how psychological and social factors influence gender biases at different stages of recovery (Beran & Scafide, 2022; Caccese, et al. 2023; CARE Consortium Investigators et al., 2018; Lempke, Oldham, et al. 2023; Sinnott et al., 2023). Researchers may want to expand on the current Rasch Partial Credit Model to account for additional parameters to understand better how much external factors influence accurate symptom reporting. Rasch modeling will be a useful tool for researchers in the concussion field to evaluate the relationship between sociological pressures, such as reporting intentions, and diagnostic measures, such as symptom presentation, on the variability of recovery length.

### Future SCAT Assessment Recommendations

A female-athlete-specific section in future versions of the SCAT is essential due to accumulating evidence that female athletes experience and report concussion symptoms differently from their male counterparts. Female athletes are more likely to report symptoms such as migraines, mood disturbances, and neck pain after a concussion, which may be linked to anatomical, hormonal, and physiological differences. These differences not only affect symptom severity but can also influence recovery duration, as female athletes often report prolonged symptom durations compared to male athletes.

The current SCAT assessment, however, remains largely agnostic to gender differences, potentially leading to under-recognition or misinterpretation of symptoms in female athletes. By incorporating a new section in future versions of the SCAT dedicated to symptoms and issues more commonly reported by female athletes, such as hormonal influences on mood and menstrual cycle irregularities, the SCAT could provide a more accurate and comprehensive picture of concussion specifically as it pertains to women. This tailored approach would support clinicians in identifying concussion effects more precisely and creating individualized care plans that consider the unique recovery patterns of female athletes. A female-athlete-specific section in the SCAT would, thus, represent a critical step forward in equitable, evidence-based concussion care for athletes across all sports and competition levels.

More specifically, to enhance the SCAT’s sensitivity to the unique experiences of female athletes with SRC, based upon the discriminant analysis performed here, five additional assessment items concering the following might be considered:

#### Menstrual Cycle Changes and Symptoms

Concussions can impact the menstrual cycle resulting in irregularities or heightened premenstrual symptoms, which, themselves, may complicate recovery. An item asking about recent changes in menstrual patterns or cycle-related symptom severity would allow clinicians to monitor potential hormonal impacts that may influence both symptoms and healing time in female athletes.

#### Mood Changes and Emotional Sensitivity

Research indicates that female athletes are more likely to report mood swings, irritability, and emotional sensitivity post-concussion. Adding an item specifically assessing mood disturbances (“Have you experienced increased mood swings, irritability, or emotional sensitivity?”) could help clinicians monitor this common symptom and inform recovery strategies.

#### Sleep Disturbances Related to Hormonal Fluctuations

Likewise, hormonal fluctuations can affect sleep quality in female athletes, which may exacerbate concussion recovery. A targeted item assessing sleep issues with attention to any recent menstrual or hormonal changes (e.g., “Have you experienced disrupted sleep, especially during your menstrual cycle?”) could give a fuller picture of factors influencing recovery.

#### Migraine-Like Symptoms

While headache is included in the SCAT, female athletes often report migraine-like symptoms, such as throbbing pain and heightened sensitivity to light or sound, more frequently than male athletes following concussion. A more specific item assessing the nature and intensity of headache symptoms (e.g., “Is the headache migraine-like, with throbbing or sensitivity to light/sound?”) could capture this experience more accurately.

#### Neck Pain and Whiplash Sensitivity

Female athletes suffer higher instances of neck pain and whiplash-like symptoms post-injury, likely due to anatomical and muscular differences (Lin et al., 2018). A new item assessing neck-related symptoms or pain more carefully would allow athletic trainers and clinicians to differentiate these from primary concussion symptoms, thereby refining diagnosis and treatment options.

Incorporating a new section including items on these topics could make future versions of the SCAT more responsive to the unique symptom experiences of female athletes, potentially leading to more personalized and effective concussion management strategies.

## Conclusion

In conclusion, this quantitatively-focused examination highlights the multidimensional nature of concussion self-reporting of symptoms as measured by the SCAT Symptom Severity Checklist and underscores the importance of 1) the multi-factorial nature of the SCAT symptom self-reporting, as well as 2) more carefully considering gender differences in concussion assessment. The findings suggest that an athlete-gender-agnostic approach to concussion symptom severity may not be appropriate and that gender-specific considerations should be integrated into clinical assessments and treatment plans. By adopting a more nuanced, multidimensional approach, athletic training staff and healthcare providers can ensure more precise diagnosis and tailored interventions, ultimately improving outcomes for all athletes. Neuropsychological testing is recommended to remain a key component in evaluating complex concussions, although it is not currently considered essential for assessing simple concussions. While the SCAT assessment and the analyses examined here significantly enhances both the understanding of concussion effects and the potential for management of individual athletes, they should not serve as the sole basis for decisions regarding time away from play or return-to-play readiness. Nevertheless, the clinical management of concussions specific to the symptoms female athletes tend to report is not often part of current SRC assessments (Malcom et al., 2023) as currently practiced, as exemplified by the SCAT. If included in future versions of the SCAT, the additions of self-report items, as recommended here, strategically added to the sub-scale structure or as its own sub-scale, altogether, would undoubtedly provide important context to individualized SRC treatment approaches.

## Data Availability

All data produced are available online at https://fitbir.nih.gov

## Acknowledgements

The authors wish to thank contributions on an earlier version of this manuscript from Ms. Sydney Cushing of the Department of Psychology at the University of Virginia.

## References

Asken, B. M., Bauer, R. M., DeKosky, S. T., Svingos, A. M., Hromas, G., Boone, J. K., DuBose, D. N., Hayes, R. L., & Clugston, J. R. (2018). Concussion BASICS III: Serum biomarker changes following sport-related concussion. Neurology, 91(23), e2133–e2143. 10.1212/WNL.0000000000006617

Bakker, L. A., Schröder, C. D., Van Es, M. A., Westers, P., Visser-Meily, J. M. A., & Van Den Berg, L. H. (2017). Assessment of the factorial validity and reliability of the ALSFRS-R: A revision of its measurement model. Journal of Neurology, 264(7), 1413–1420. 10.1007/s00415-017-8538-4

Begasse de Dhaem, O., Barr, W. B., Balcer, L. J., Galetta, S. L., & Minen, M. T. (2017). Post-traumatic headache: The use of the sport concussion assessment tool (SCAT-3) as a predictor of post-concussion recovery. The Journal of Headache and Pain, 18(1), 60. 10.1186/s10194-017-0767-5

Benjamini, Y., & Hochberg, Y. (1995). Controlling the False Discovery Rate: A Practical and Powerful Approach to Multiple Testing. Journal of the Royal Statistical Society: Series B (Methodological), 57(1), 289–300. 10.1111/j.2517-6161.1995.tb02031.x

Beran, K. M., & Scafide, K. N. (2022). Factors Related to Concussion Knowledge, Attitudes, and Reporting Behaviors in US High School Athletes: A Systematic Review. Journal of School Health, 92(4), 406–417. 10.1111/josh.13140

Bollmann, S., Berger, M., & Tutz, G. (2018). Item-Focused Trees for the Detection of Differential Item Functioning in Partial Credit Models. Educational and Psychological Measurement, 78(5), 781–804. 10.1177/0013164417722179

Brett, B. L., Kramer, M. D., McCrea, M. A., Broglio, S. P., McAllister, T. W., Nelson, L. D., the CARE Consortium Investigators, Hazzard, J. B., Kelly, L. A., Ortega, J., Port, N., Pasquina, P. F., Jackson, J., Cameron, K. L., Houston, M. N., Goldman, J. T., Giza, C., Buckley, T., Clugston, J. R., … Susmarski, A. (2020). Bifactor Model of the Sport Concussion Assessment Tool Symptom Checklist: Replication and Invariance Across Time in the CARE Consortium Sample. The American Journal of Sports Medicine, 48(11), 2783–2795. 10.1177/0363546520946056

Broglio, S. P., Cantu, R. C., Gioia, G. A., Guskiewicz, K. M., Kutcher, J., Palm, M., Valovich McLeod, T. C., & National Athletic Trainer’s Association (2014). National Athletic Trainers’ Association position statement: management of sport concussion. Journal of athletic training, 49(2), 245–265. 10.4085/1062-6050-49.1.07

Broshek, D. K., Kaushik, T., Freeman, J. R., Erlanger, D., Webbe, F., & Barth, J. T. (2005). Gender differences in outcome following sports-related concussion. Journal of Neurosurgery, 102(5), 856–863. 10.3171/jns.2005.102.5.0856

Brown, D. A., Elsass, J. A., Miller, A. J., Reed, L. E., & Reneker, J. C. (2015a). Differences in Symptom Reporting Between Males and Females at Baseline and After a Sports-Related Concussion: A Systematic Review and Meta-Analysis. Sports Medicine, 45(7), 1027–1040. 10.1007/s40279-015-0335-6

Buchardt, A-S., Christensen, K. B., & Jensen, S. N. (2023). Visualizing Rasch item fit using conditional item characteristic curves in R. Psychological Test and Assessment Modeling, 2, 206.

Cantu, R. C., & Register-Mihalik, J. K. (2011). Considerations for Return-to-Play and Retirement Decisions After Concussion. PM&R, 3(10S2). 10.1016/j.pmrj.2011.07.013

Colins, O. F., Noom, M., & Vanderplasschen, W. (2012). Youth Psychopathic Traits Inventory-Short Version: A Further Test of the Internal Consistency and Criterion Validity. Journal of Psychopathology and Behavioral Assessment, 34(4), 476–486. 10.1007/s10862-012-9299-0

CARE Consortium Investigators, Broglio, S. P., Katz, B. P., Zhao, S., McCrea, M., & McAllister, T. (2018). Test-Retest Reliability and Interpretation of Common Concussion Assessment Tools: Findings from the NCAA-DoD CARE Consortium. Sports Medicine, 48(5), 1255–1268. 10.1007/s40279-017-0813-0

Chandran, A., Boltz, A. J., Morris, S. N., Robison, H. J., Nedimyer, A. K., Collins, C. L., & Register-Mihalik, J. K. (2022). Epidemiology of Concussions in National Collegiate Athletic Association (NCAA) Sports: 2014/15-2018/19. The American Journal of Sports Medicine, 50(2), 526–536. 10.1177/03635465211060340

Chin, E. Y., Nelson, L. D., Barr, W. B., McCrory, P., & McCrea, M. A. (2016). Reliability and Validity of the Sport Concussion Assessment Tool–3 (SCAT3) in High School and Collegiate Athletes. The American Journal of Sports Medicine, 44(9), 2276–2285. 10.1177/0363546516648141

Choi, S. W., Schalet, B., Cook, K. F., & Cella, D. (2014). Establishing a common metric for depressive symptoms: linking the BDI-II, CES-D, and PHQ-9 to PROMIS depression. Psychological assessment, 26(2), 513–527. 10.1037/a0035768

Covassin, T., & Elbin, R. J. (2011). The Female Athlete: The Role of Gender in the Assessment and Management of Sport-Related Concussion. Clinics in Sports Medicine, 30(1), 125–131. 10.1016/j.csm.2010.08.001

Covassin, T., Elbin, R., Kontos, A., & Larson, E. (2010). Investigating baseline neurocognitive performance between male and female athletes with a history of multiple concussion. Journal of Neurology, Neurosurgery & Psychiatry, 81(6), 597–601. 10.1136/jnnp.2009.193797

Covassin, T., Swanik, C. B., Sachs, M., Kendrick, Z., Schatz, P., Zillmer, E., Kaminaris, C., Iverson, G., & Stearne, D. J. (2006). Gender differences in baseline neuropsychological function and concussion symptoms of collegiate athletes * Commentary * Commentary. British Journal of Sports Medicine, 40(11), 923–927. 10.1136/bjsm.2006.029496

Covassin, T., Swanik, C. B., & Sachs, M. L. (2003). Gender Differences and the Incidence of Concussions Among Collegiate Athletes. Journal of Athletic Training, 38(3), 238–244.

de Ayala, R. J. (2013). Theory and Practice of Item Response Theory. Guilford Publications.

Dick, R. W. (2009). Is there a gender difference in concussion incidence and outcomes? British Journal of Sports Medicine, 43(Suppl_1), i46–i50. 10.1136/bjsm.2009.058172

D’Lauro, C., Jones, E. R., Swope, L. M., Anderson, M. N., Broglio, S., & Schmidt, J. D. (2022). Under-representation of female athletes in research informing influential concussion consensus and position statements: An evidence review and synthesis. British Journal of Sports Medicine, 56(17), 981–987. 10.1136/bjsports-2021-105045

Dorans, N. J., & Holland, P. W. (1992). DIF DETECTION AND DESCRIPTION: MANTEL-HAENSZEL AND STANDARDIZATION1,2. ETS Research Report Series, 1992(1). 10.1002/j.2333-8504.1992.tb01440.x

Edelstein, R., & Van Horn, J.D. (2023). Modulating Factors Affecting Sports-Related Concussion Exposures: A Systematic Review and Analysis. medRxiv, DOI:10.1101/2023.03.08.23286974.

Edelstein, R., Gutterman, S., Newman, B., & Van Horn, J. D. (2024). Assessment of Sports Concussion in Female Athletes: A Role for Neuroinformatics?. Neuroinformatics, 22(4), 607–618. 10.1007/s12021-024-09680-8

Embretson SE, Reise SP. 2000. Item Response Theory for Psychologists. Mahwah, NJ: Erlbaum

Echemendia, R. J., Meeuwisse, W., McCrory, P., Davis, G. A., Putukian, M., Leddy, J., Makdissi, M., Sullivan, S. J., Broglio, S. P., Raftery, M., Schneider, K., Kissick, J., McCrea, M., Dvorak, J., Sills, A. K., Aubry, M., Engebretsen, L., Loosemore, M., Fuller, G., … Herring, S. (2017). The Sport Concussion Assessment Tool 5th Edition (SCAT5). British Journal of Sports Medicine, bjsports-2017-097506. 10.1136/bjsports-2017-097506

Garcia, G.-G. P., Yang, J., Lavieri, M. S., McAllister, T. W., McCrea, M. A., Broglio, S. P., & on behalf of the CARE Consortium Investigators. (2020). Optimizing Components of the Sport Concussion Assessment Tool for Acute Concussion Assessment. Neurosurgery, 87(5), 971–981. 10.1093/neuros/nyaa150

Gessel, L. M., Fields, S. K., Collins, C. L., Dick, R. W., & Comstock, R. D. (2007). Concussions among United States high school and collegiate athletes. Journal of Athletic Training, 42(4), 495–503.

Granito, V. J. (2002). Psychological response to athletic injury: Gender differences. Journal of Sport Behavior, 25(3), 243+. Gale Academic OneFile.

Golino, H. F., & Epskamp, S. (2017). Exploratory graph analysis: A new approach for estimating the number of dimensions in psychological research. PLOS ONE, 12(6), e0174035. 10.1371/journal.pone.0174035

Golino, H., Shi, D., Christensen, A. P., Garrido, L. E., Nieto, M. D., Sadana, R., Thiyagarajan, J. A., & Martinez-Molina, A. (2020). Investigating the performance of exploratory graph analysis and traditional techniques to identify the number of latent factors: A simulation and tutorial. Psychological methods, 25(3), 292–320. 10.1037/met0000255

Guskiewicz, K. M., Register-Mihalik, J., McCrory, P., McCrea, M., Johnston, K., Makdissi, M., Dvořák, J., Davis, G., & Meeuwisse, W. (2013). Evidence-based approach to revising the SCAT2: Introducing the SCAT3: Table 1. British Journal of Sports Medicine, 47(5), 289–293. 10.1136/bjsports-2013-092225

Hamel, J.-F., Sébille, V., Challet-Bouju, G., & Hardouin, J.-B. (2016). Partial Credit Model: Estimations and Tests of Fit with Pcmodel. The Stata Journal: Promoting Communications on Statistics and Stata, 16(2), 464–481. 10.1177/1536867X1601600212

Harmon, K. G., Drezner, J. A., Gammons, M., Guskiewicz, K. M., Halstead, M., Herring, S. A., Kutcher, J. S., Pana, A., Putukian, M., & Roberts, W. O. (2013). American Medical Society for Sports Medicine position statement: Concussion in sport. British Journal of Sports Medicine, 47(1), 15–26. 10.1136/bjsports-2012-091941

Heck, S. J., Acord-Vira, A., & Davis, D. R. (2023). Gender differences in college students’ knowledge of concussion and concussion education sources. Concussion, 8(3), CNC108. 10.2217/cnc-2023-0001

Kieffer, E. E., Brolinson, P. G., Maerlender, A. E., Smith, E. P., & Rowson, S. (2021). In-Season Concussion Symptom Reporting in Male and Female Collegiate Rugby Athletes. Neurotrauma Reports, 2(1), 503–511. 10.1089/neur.2021.0050

Klecka, W. R. (1980). Discriminant Analysis. Beverly Hills, CA: Sage.

Kochick, V., Sinnott, A. M., Eagle, S. R., Bricker, I. R., Collins, M. W., Mucha, A., Connaboy, C., & Kontos, A. P. (2022). The Dynamic Exertion Test for Sport-Related Concussion: A Comparison of Athletes at Return-to-Play and Healthy Controls. International Journal of Sports Physiology and Performance, 17(6), 834–843. 10.1123/ijspp.2021-0258

Kolen, M. J., & Brennan, R. L. (2004). Test equating, scaling, and linking: Methods and practices. Springer. Retrieved from https://books.google.com/books?id=tusOwjb7LWsC

Kroshus, E., Lowry, S. J., Garrett, K., Hays, R., Hunt, T., & Chrisman, S. P. D. (2021a). Development of a scale to measure expected concussion reporting behavior. Injury Epidemiology, 8(1), 70. 10.1186/s40621-021-00364-4

Langer, M. M., Hill, C. D., Thissen, D., Burwinkle, T. M., Varni, J. W., & DeWalt, D. A. (2008a). Item response theory detected differential item functioning between healthy and ill children in quality-of-life measures. Journal of Clinical Epidemiology, 61(3), 268–276. 10.1016/j.jclinepi.2007.05.002

Langlois, J. A., Rutland-Brown, W., & Wald, M. M. (2006). The Epidemiology and Impact of Traumatic Brain Injury: A Brief Overview. Journal of Head Trauma Rehabilitation, 21(5), 375–378. 10.1097/00001199-200609000-00001

Lin, C. Y., Casey, E., Herman, D. C., Katz, N., & Tenforde, A. S. (2018). Gender Differences in Common Sports Injuries. PM&R, 10(10), 1073–1082. 10.1016/j.pmrj.2018.03.008

Linacre, J. (2023). WINSTEPS®. https://www.winsteps.com/index.htm

Lempke, L. B., Caccese, J. B., Syrydiuk, R. A., Buckley, T. A., Chrisman, S. P. D., Clugston, J. R., Eckner, J. T., Ermer, E., Esopenko, C., Jain, D., Kelly, L. A., Memmini, A. K., Mozel, A. E., Putukian, M., Susmarski, A., Pasquina, P. F., McCrea, M. A., McAllister, T. W., Broglio, S. P., … CARE Consortium Investigators. (2023). Female Collegiate Athletes’ Concussion Characteristics and Recovery Patterns: A Report from the NCAA-DoD CARE Consortium. Annals of Biomedical Engineering. 10.1007/s10439-023-03367-y

Lempke, L. B., Johnson, R. S., Schmidt, J. D., & Lynall, R. C. (2020). Clinical versus Functional Reaction Time: Implications for Postconcussion Management. Medicine & Science in Sports & Exercise, 52(8), 1650–1657. 10.1249/MSS.0000000000002300

Lempke, L. B., Oldham, J. R., Passalugo, S., Willwerth, S. B., Berkstresser, B., Wang, F., Howell, D. R., & Meehan, W. P. (2023). Influential Factors and Preliminary Reference Data for a Clinically Feasible, Functional Reaction Time Assessment: The Standardized Assessment of Reaction Time. Journal of Athletic Training, 58(2), 112–119. 10.4085/1062-6050-0073.22

Lempke, L. B., Passalugo, S., Baranker, B. T., Hunt, D., Berkstresser, B., Wang, F., Meehan, W. P., & Howell, D. R. (2022). Relationship and Latent Factors Between Clinical Concussion Assessments and the Functional Standardized Assessment of Reaction Time (StART). Clinical Journal of Sport Medicine, 32(6), e591–e597. 10.1097/JSM.0000000000001061

Lempke, L. B., Shumski, E. J., Prato, T. A., & Lynall, R. C. (2023). Reliability and Minimal Detectable Change of the Standardized Assessment of Reaction Time. Journal of Athletic Training, 58(6), 579–587. 10.4085/1062-6050-0391.22

Malcolm, D. (2023). Some problems of research exploring gender differences in sport-related concussions: A narrative review. Research in Sports Medicine, 1–10. 10.1080/15438627.2023.2271604

Masters, G. N., & Wright, B. D. (1997). The Partial Credit Model. In W. J. Van Der Linden & R. K. Hambleton (Eds.), Handbook of Modern Item Response Theory (pp. 101–121). Springer New York. 10.1007/978-1-4757-2691-6_6

Masters, G.N. (1982). A Rasch model for partial credit scoring. Psychometrika, 47, 149–174.

McCrory, P., Meeuwisse, W., Aubry, M., Cantu, B., Dvořák, J., Echemendia, R., Engebretsen, L., Johnston, K., Kutcher, J., Raftery, M., Sills, A., Benson, B., Davis, G., Ellenbogen, R., Guskiewicz, K., Herring, S. A., Iverson, G., Jordan, B., Kissick, J., … Turner, M. (2013a). Consensus statement on Concussion in Sport – The 4th International Conference on Concussion in Sport held in Zurich, November 2012. Physical Therapy in Sport, 14(2), e1–e13. 10.1016/j.ptsp.2013.03.002

McCrory, P., Meeuwisse, W., Johnston, K., Dvorak, J., Aubry, M., Molloy, M., & Cantu, R. (2009). Consensus Statement on Concussion in Sport 3rd International Conference on Concussion in Sport Held in Zurich, November 2008. Clinical Journal of Sport Medicine, 19(3), 185–200. 10.1097/JSM.0b013e3181a501db

Meehan, W. P., Mannix, R. C., O’Brien, M. J., & Collins, M. W. (2013). The Prevalence of Undiagnosed Concussions in Athletes. Clinical Journal of Sport Medicine, 23(5), 339–342. 10.1097/JSM.0b013e318291d3b3

Mihalik, J. P., Ondrak, K. S., Guskiewicz, K. M., & McMurray, R. G. (2009). The effects of menstrual cycle phase on clinical measures of concussion in healthy college-aged females. Journal of Science and Medicine in Sport, 12(3), 383–387. 10.1016/j.jsams.2008.05.003

National Research Council (U.S.), Graham, R., Rivara, F. P., Ford, M. A., Spicer, C. M., & Institute of Medicine (U.S.) (Eds.). (2014). Sports-related concussions in youth: Improving the science, changing the culture. The National Academies Press.

Nelson, L. D., Kramer, M. D., Patrick, C. J., & McCrea, M. A. (2018). Modeling the Structure of Acute Sport-Related Concussion Symptoms: A Bifactor Approach. Journal of the International Neuropsychological Society, 24(8), 793–804. 10.1017/S1355617718000462

O’Connor, K. L., Baker, M. M., Dalton, S. L., Dompier, T. P., Broglio, S. P., & Kerr, Z. Y. (2017). Epidemiology of Sport-Related Concussions in High School Athletes: National Athletic Treatment, Injury and Outcomes Network (NATION), 2011–2012 Through 2013–2014. Journal of Athletic Training, 52(3), 175–185. 10.4085/1062-6050-52.1.15

Pearson, K. (1901). On lines and planes of closest fit to systems of points in space. Philosophical Magazine, 2(11), 559–572

Preiss-Farzanegan, S. J., Chapman, B., Wong, T. M., Wu, J., & Bazarian, J. J. (2009). The Relationship Between Gender and Postconcussion Symptoms After Sport-Related Mild Traumatic Brain Injury. PM&R, 1(3), 245–253. 10.1016/j.pmrj.2009.01.011

Randolph, C., Millis, S., Barr, W. B., McCrea, M., Guskiewicz, K. M., Hammeke, T. A., & Kelly, J. P. (2009a). Concussion Symptom Inventory: An Empirically Derived Scale for Monitoring Resolution of Symptoms Following Sport-Related Concussion. Archives of Clinical Neuropsychology, 24(3), 219–229. 10.1093/arclin/acp025

Rencher, A. C. (1992). Interpretation of Canonical Discriminant Functions, Canonical Variates, and Principal Components. The American Statistician, 46(3), 217–225. 10.1080/00031305.1992.10475889

Rutherford, WilliamH., Merrett, JohnD., & Mcdonali, JohnR. (1977). SEQUELÆ OF CONCUSSION CAUSED BY MINOR HEAD INJURIES. The Lancet, 309(8001), 1–4. 10.1016/S0140-6736(77)91649-X

Sport Concussion Assessment Tool 6 (SCAT6). (2023). British Journal of Sports Medicine, 57(11), 622–631. 10.1136/bjsports-2023-107036

Schatz, P., Moser, R. S., Covassin, T., & Karpf, R. (2011). Early Indicators of Enduring Symptoms in High School Athletes With Multiple Previous Concussions. Neurosurgery, 68(6), 1562–1567. 10.1227/NEU.0b013e31820e382e

Sinnott, A. M., Eagle, S. R., Kochick, V., Bricker, I. R., Collins, M. W., Sparto, P. J., Flanagan, S. D., Elbin, R. J., Connaboy, C., & Kontos, A. P. (2023). Test-Retest, Interrater Reliability, and Minimal Detectable Change of the Dynamic Exertion Test (EXiT) for Concussion. Sports Health: A Multidisciplinary Approach, 15(3), 410–421. 10.1177/19417381221093556

Spearman, C. (1904). “General Intelligence,” objectively determined and measured. American Journal of Psychology, 15(2), 201–292.

Stemper, B. D. (2022). Sport-related concussion: The role of repetitive head impact exposure. In Cellular, Molecular, Physiological, and Behavioral Aspects of Traumatic Brain Injury (pp. 29–40). Elsevier. 10.1016/B978-0-12-823036-7.00023-2

Tennant, A., & Conaghan, P. G. (2007). The Rasch measurement model in rheumatology: What is it and why use it? When should it be applied, and what should one look for in a Rasch paper? Arthritis Care & Research, 57(8), 1358–1362. 10.1002/art.23108

the EORTC Quality of Life Group and the Quality of Life Cross-Cultural Meta-Analysis Group, Scott, N. W., Fayers, P. M., Aaronson, N. K., Bottomley, A., De Graeff, A., Groenvold, M., Gundy, C., Koller, M., Petersen, M. A., & Sprangers, M. A. (2010a). Differential item functioning (DIF) analyses of health-related quality of life instruments using logistic regression. Health and Quality of Life Outcomes, 8(1), 81. 10.1186/1477-7525-8-81

Wallace, J., Covassin, T., & Beidler, E. (2017a). Gender Differences in High School Athletes’ Knowledge of Sport-Related Concussion Symptoms and Reporting Behaviors. Journal of Athletic Training, 52(7), 682– 688. 10.4085/1062-6050-52.3.06

Warmath, D., & Winterstein, A. P. (2020). A Social-Marketing Intervention and Concussion-Reporting Beliefs. Journal of athletic training, 55(10), 1035–1045. 10.4085/1062-6050-242-19

Wilmoth, K., Magnus, B. E., McCrea, M. A., & Nelson, L. D. (2020). Preliminary Validation of an Abbreviated Acute Concussion Symptom Checklist Using Item Response Theory. The American Journal of Sports Medicine, 48(12), 3087–3093. 10.1177/0363546520953440

Xanthopoulos, P., Pardalos, P. M., & Trafalis, T. B. (2013). Linear Discriminant Analysis. In P. Xanthopoulos, P. M. Pardalos, & T. B. Trafalis, Robust Data Mining (pp. 27–33). Springer New York. 10.1007/978-1-4419-9878-1_4

Yengo-Kahn, A. M., Hale, A. T., Zalneraitis, B. H., Zuckerman, S. L., Sills, A. K., & Solomon, G. S. (2016). The Sport Concussion Assessment Tool: A systematic review. Neurosurgical Focus, 40(4), E6. 10.3171/2016.1.FOCUS15611

Zuckerman, S. L., Apple, R. P., Odom, M. J., Lee, Y. M., Solomon, G. S., & Sills, A. K. (2014). Effect of gender on symptoms and return to baseline in sport-related concussion: Clinical article. Journal of Neurosurgery: Pediatrics, 13(1), 72–81. 10.3171/2013.9.PEDS13257

Zuckerman, S. L., Kerr, Z. Y., Yengo-Kahn, A., Wasserman, E., Covassin, T., & Solomon, G. S. (2015). Epidemiology of Sports-Related Concussion in NCAA Athletes From 2009-2010 to 2013-2014: Incidence, Recurrence, and Mechanisms. The American Journal of Sports Medicine, 43(11), 2654–2662. 10.1177/0363546515599634

